# Optimal policy determination in sequential systemic and locoregional therapy of oropharyngeal squamous carcinomas: A patient-physician digital twin dyad with deep Q-learning for treatment selection

**DOI:** 10.1101/2021.04.07.21255092

**Authors:** Elisa Tardini, Xinhua Zhang, Guadalupe Canahuate, Andrew Wentzel, Abdallah S. R. Mohamed, Lisanne Van Dijk, Clifton D. Fuller, G. Elisabeta Marai

**Author notes:** ***Co-corresponding authors:*** Correspondence to Elisa Tardini and Dr. Clifton D. Fuller. Contributing authors.

## Abstract

**Purpose:** Currently, selection of patients for sequential vs. concurrent chemotherapy/radiation regimens lacks evidentiary support, and it is based on locally-optimal decisions for each step. We aim to optimize the multi-step treatment of head and neck cancer patients and to predict multiple patient survival and toxicity outcomes, and we develop, apply, and evaluate a first application of deep-Q-learning (DQL) and simulation to this problem.

**Patients and methods:** The treatment decision DQL digital twin and the patient’s digital twin were created, trained and evaluated on a dataset of 536 oropharyngeal squamous cell carcinoma (OPC) patients with the goal of, respectively, determining the optimal treatment decisions with respect to survival and toxicity metrics, and predicting the outcomes of the optimal treatment on the patient. The models were trained on a subset of 402 patients (split randomly) and evaluated on a separate set of 134 patients. Training and evaluation of the digital twin dyad was completed in August 2020. The dataset includes 3-step sequential treatment decisions and complete relevant history of the patients cohort treated at MD Anderson Cancer Center between 2005 and 2013, with radiomics analysis performed for the segmented primary tumor volumes.

**Results:** On the validation set, 87.09% mean and 90.85% median accuracy in treatment outcome prediction, matching the clinicians’ outcomes and improving (predicted) survival rate by +3.73% (95% CI: [-0.75%, +8.96%]), and dysphagia rate by +0.75% (CI: [-4.48%, +6.72%]) when following DQL treatment decisions.

**Conclusion:** Given the prediction accuracy and predicted improvement on medically relevant outcomes yielded by this approach, this *digital twin dyad* of the patient-physician dynamic treatment problem has the potential of aiding physicians in determining the optimal course of treatment and in assessing its outcomes.

## Introduction

Head and neck cancer, which includes cancers of the larynx, throat, lips, mouth, nose, and salivary glands, is now an epidemic with 65,000 new cases in the US annually [1], whose treatment is, as in many other types of cancers, a dynamic and complex process. This therapy process involves making multiple, patient-specific treatment decisions, to maximize efficacy---e.g., reduction in tumor size, time of local region control, and survival time, while minimizing side effects [2, 3, 4]. For example, a specific patient may undergo radiotherapy alone (RT), radiotherapy with concurrent chemotherapy (CC), or induction chemotherapy (IC) [5]. After each round of IC, a decision must be made whether or not to continue IC or to start either RT or CC. These decisions are currently taken by clinician or multidisciplinary tumor boards based on pre-therapy patient characteristics or crude heuristics. Notably, current risk prediction models incorporated (e.g. AJCC Staging) in clinical decision support systems do not, by themselves, systematically direct clinicians to select an appropriate treatment that incorporates *both* oncologic and toxicity endpoints.

Further, disposition to initial induction chemotherapy is then followed by a second responsive disposition to either radiotherapy or concurrent chemoradiotherapy. Inferring the optimal treatment policies for multi-stage decisions (e.g., which treatment to give initially and then after observing treatment response, Figure 1) *post hoc* is challenging, as an optimal therapy sequence cannot be readily “pieced together” from several single-stage decisions.

**Figure 1.**
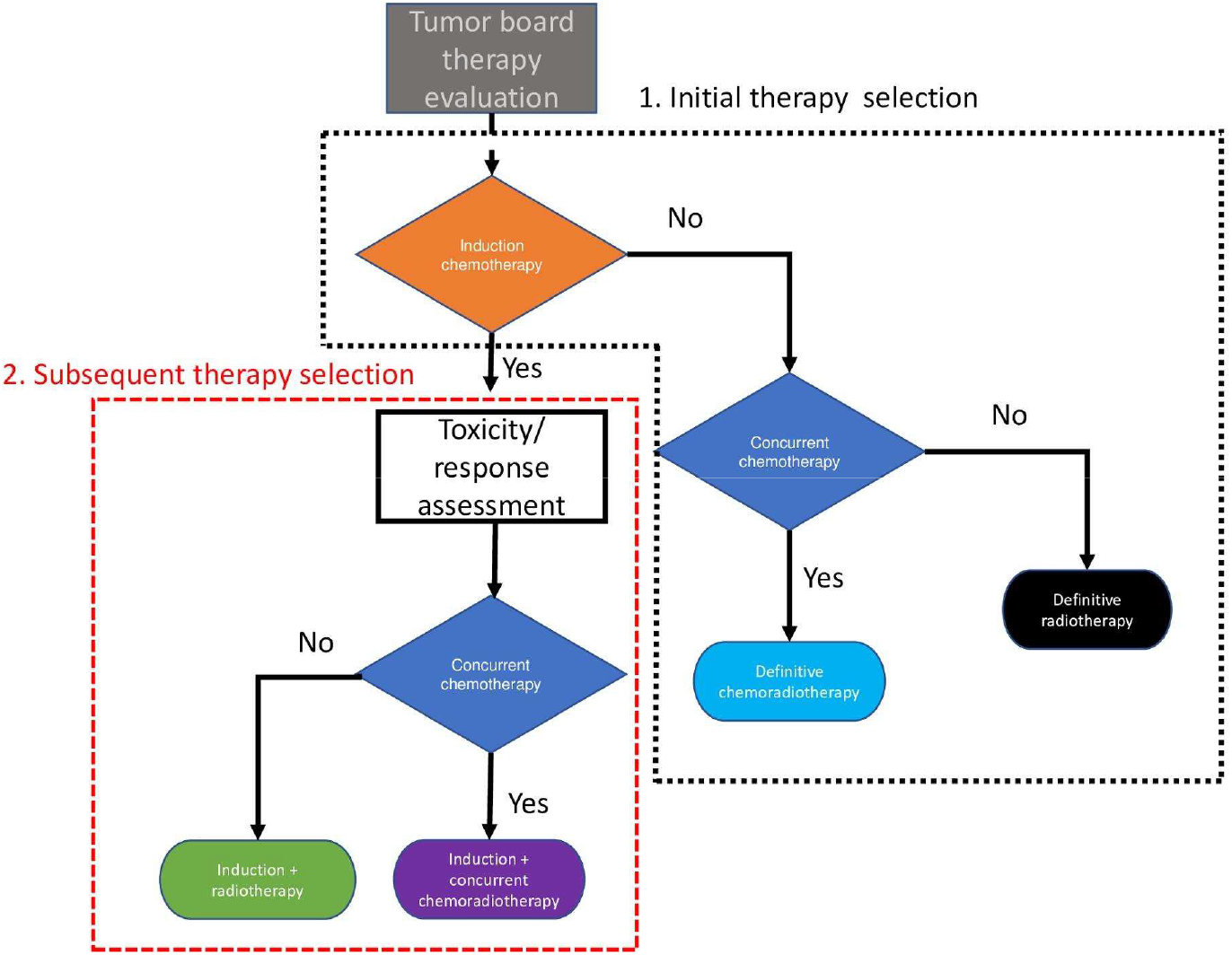
Overview of the therapy selection process. The therapy selection process shows two distinct phases: initial therapeutic selection and subsequent therapeutic selection.

For this reason, in the absence of rigorous clinical trials comparing adaptive induction chemotherapy permutations with concurrent radiotherapy, group comparison is exceedingly difficult, as simple models that account for confounders at initial disposition (e.g. propensity scores) are unequipped to incorporate sequential decision processes (e.g. the choice for chemotherapy concurrently *after* induction).

To address multi-stage models of therapy selection that incorporate both relevant cancer and side-effect considerations, we introduce an approach based on *digital twinning*, a new concept adapted to health research from the industrial world, where a digital replica (“digital twin”) of a physical entity or process is virtually recreated, with similar elements and dynamics, to perform real-time optimization and testing [6]. In healthcare, coupled digital twins, i.e., *digital twin dyads*, could be created for both patients and for the therapy process, and used to inform in a quantitative manner adaptive therapy decision-making, and to allow personalization and optimization of health outcomes, prediction and prevention of adverse events, and planning interventions [7]. By leveraging a large number of head and neck cancer cases collected at a single institutional head and neck data tumor board at the M.D. Anderson Cancer Center (MDACC), we propose a methodology approach to leverage deep Q-learning as a method to construct a *digital twin dyad* for simulation of therapy outcomes, and potential implementation as a clinical decision aid. Q-learning is a recently developed machine learning method for supervised variable selection and weighting accounting for iterative processes [8].

In this paper, we apply for the first time Q-learning methodology to dynamically select treatment, based on multiple clinically relevant outcomes from head and neck cancer patient-specific data. We use these methods to construct and develop optimal dynamic treatment strategies, i.e., digital twins of the therapy process. In conjunction with simulation models of patient data, the treatment prescription models form a patient-physician (prescriber) *digital twin dyad*, in which the prescription models act as a physician avatar, by identifying the optimal treatment for the patient, whereas the simulation models represent the patient by predicting the outcome of the treatment sequence. We evaluate the results of this digital twin dyad approach on a curated dataset of head and neck cancer patients.

## Methods

We use Deep Q-learning (DQL) [8, 9, 10, 33] to find a treatment policy that maximizes a linear combination of multiple patient outcomes, e.g., toxicological and survival outcomes. We consider three treatment decision points for each patient: (D1): *Induction Chemotherapy (IC) or Not*; (D2): *Concurrent Chemotherapy (CC) or Radiotherapy alone (RT)*; and (D3): *Neck Dissection (ND)*.

### Patient Dataset

We performed an IRB-approved, retrospective review of 536 patients with oropharyngeal squamous cell carcinoma (OPC) who were treated at MDACC between 2005 and 2013 (Table 1, and eTable 1 in Appendix). Radiomics analysis was performed [11, 12] for the segmented [13, 14] primary tumor volumes. Only patients with a minimum follow-up of 4 years, or who died before 4 years, were included in the dataset. We focus on two scenarios: a) *Overall Survival (4 Years)* (OS) as a single binary dichotomized outcome measure (i.e. the patient survived for at least 4 years after the treatment ended, coded as 1, all other events coded 0); and b) The combination of *Overall Survival (4 years)* and *Dysphagia (DP)* as a multi-outcome measure. *Dysphagia* is defined as either *Feeding Tube (FT)* or *Aspiration Rate (AR)* 6 months after the end of treatment [15, 16]. The combined outcome measure is computed only at the final step using the following formula:

**Table 1:**
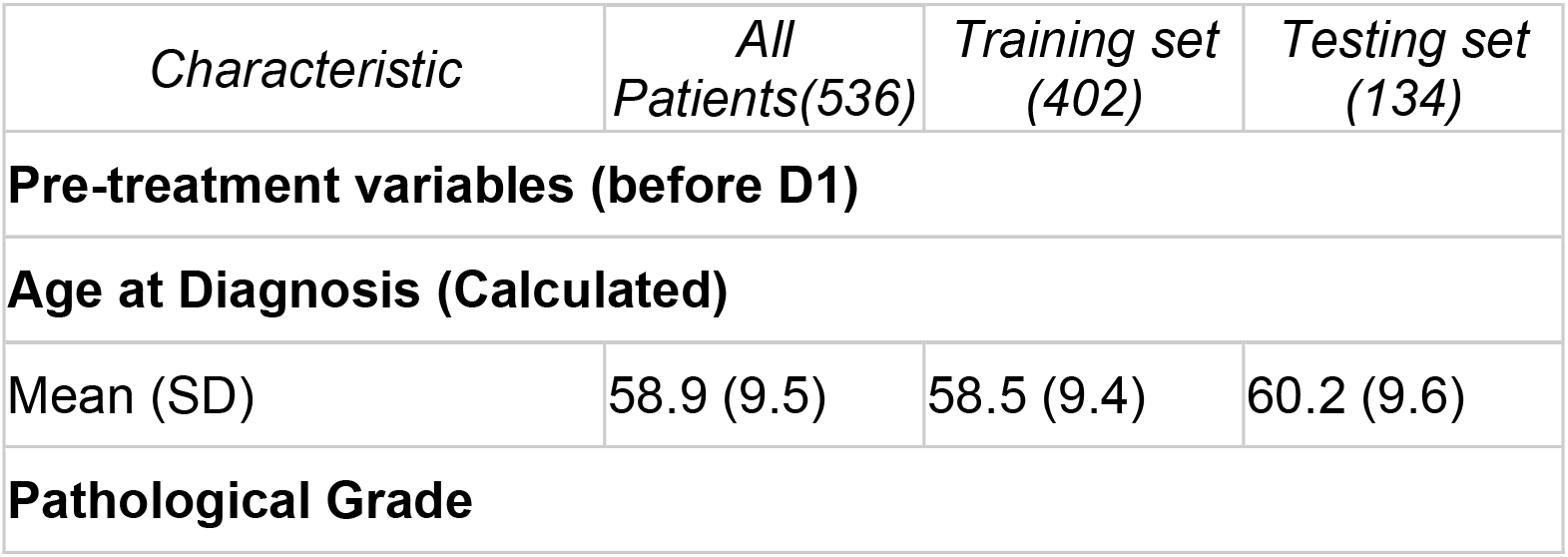
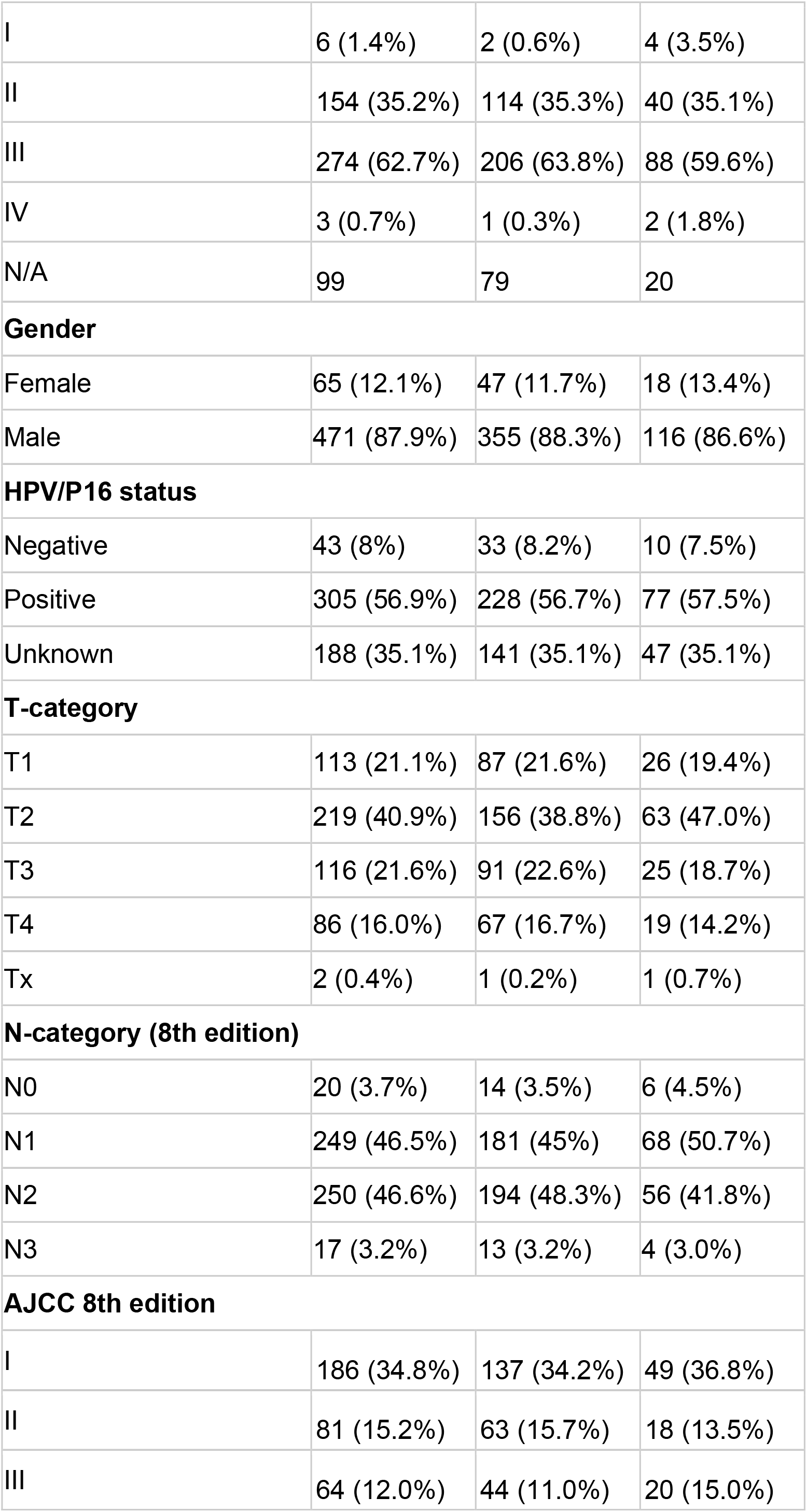
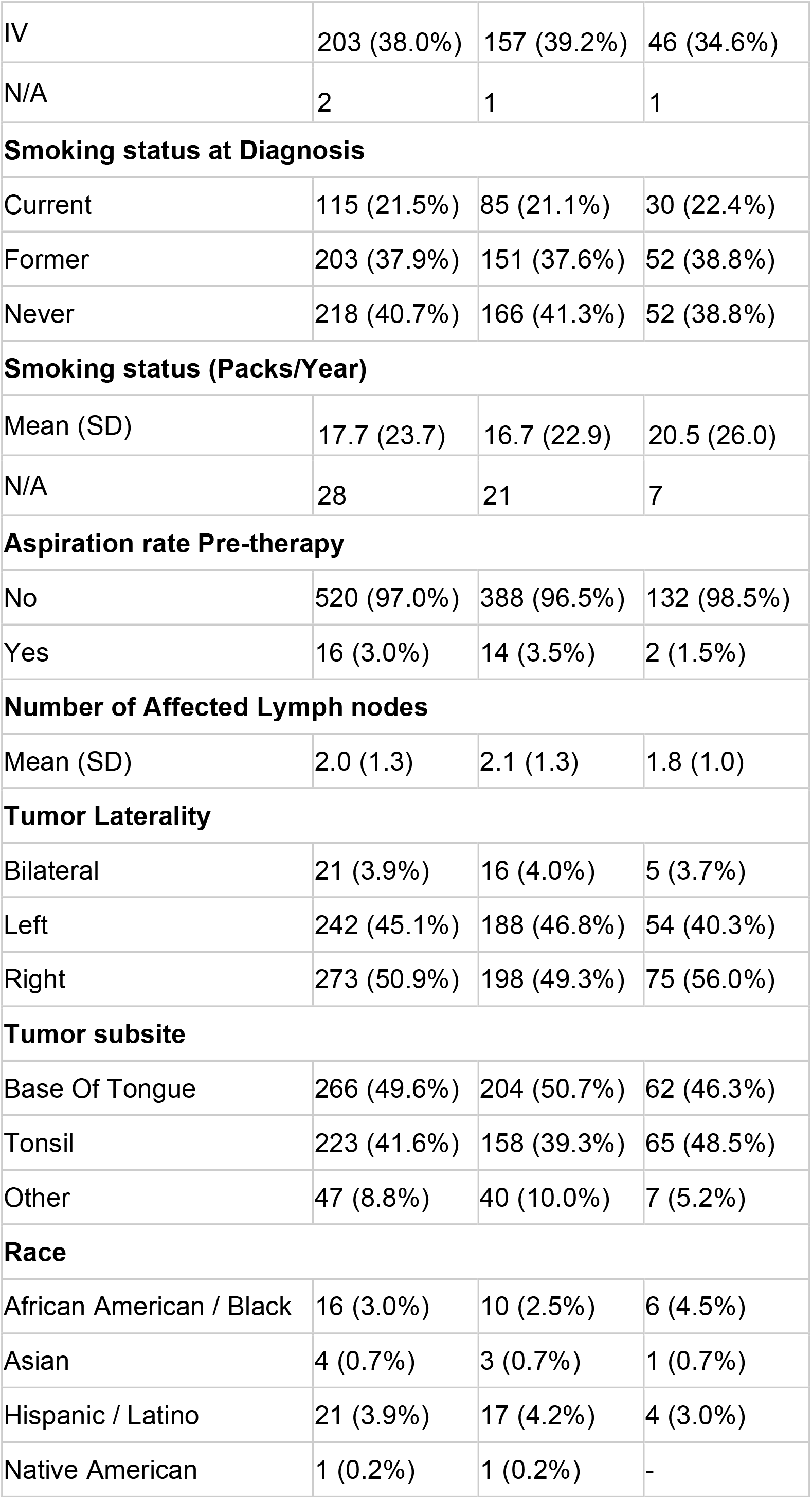
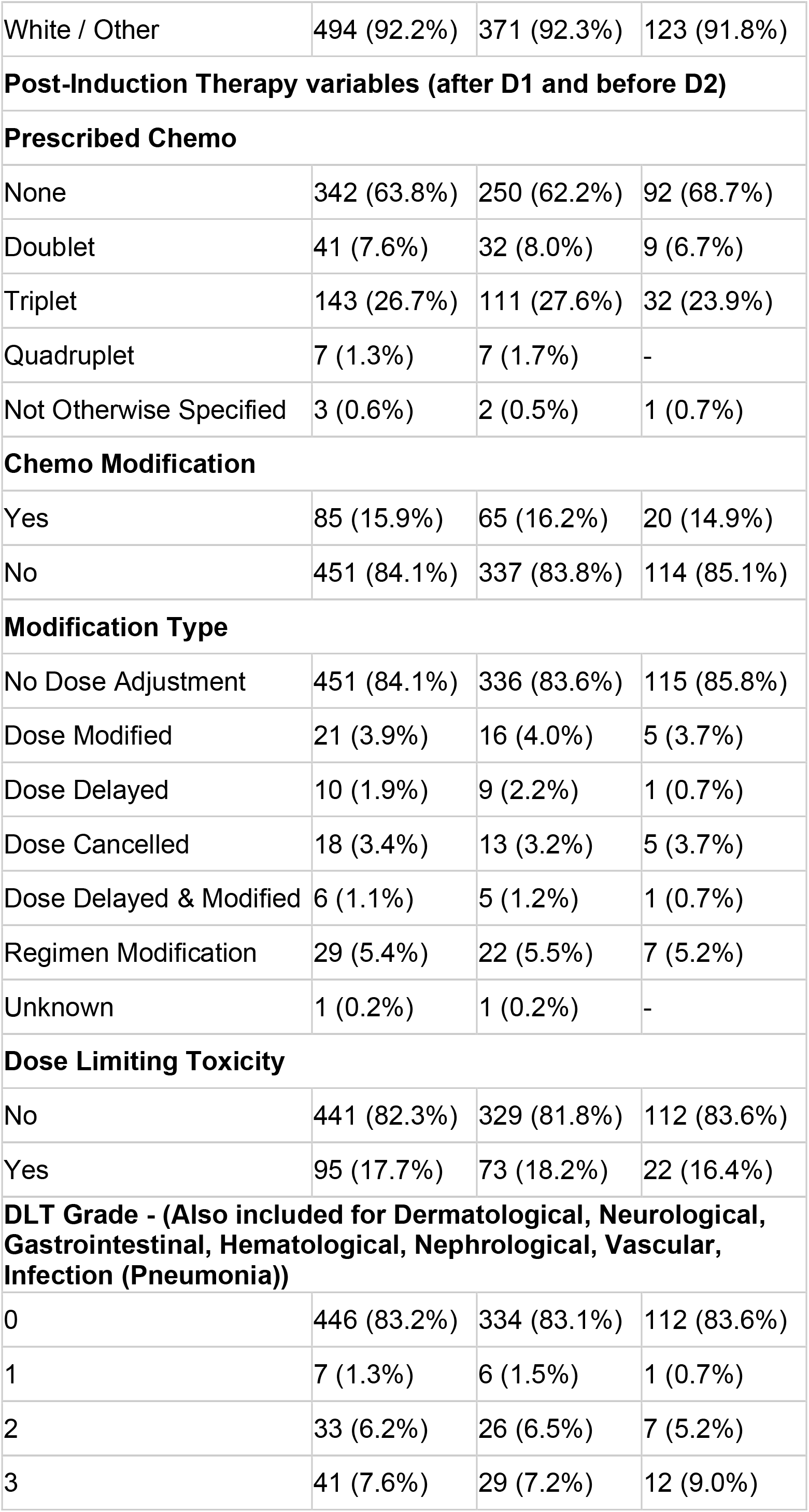
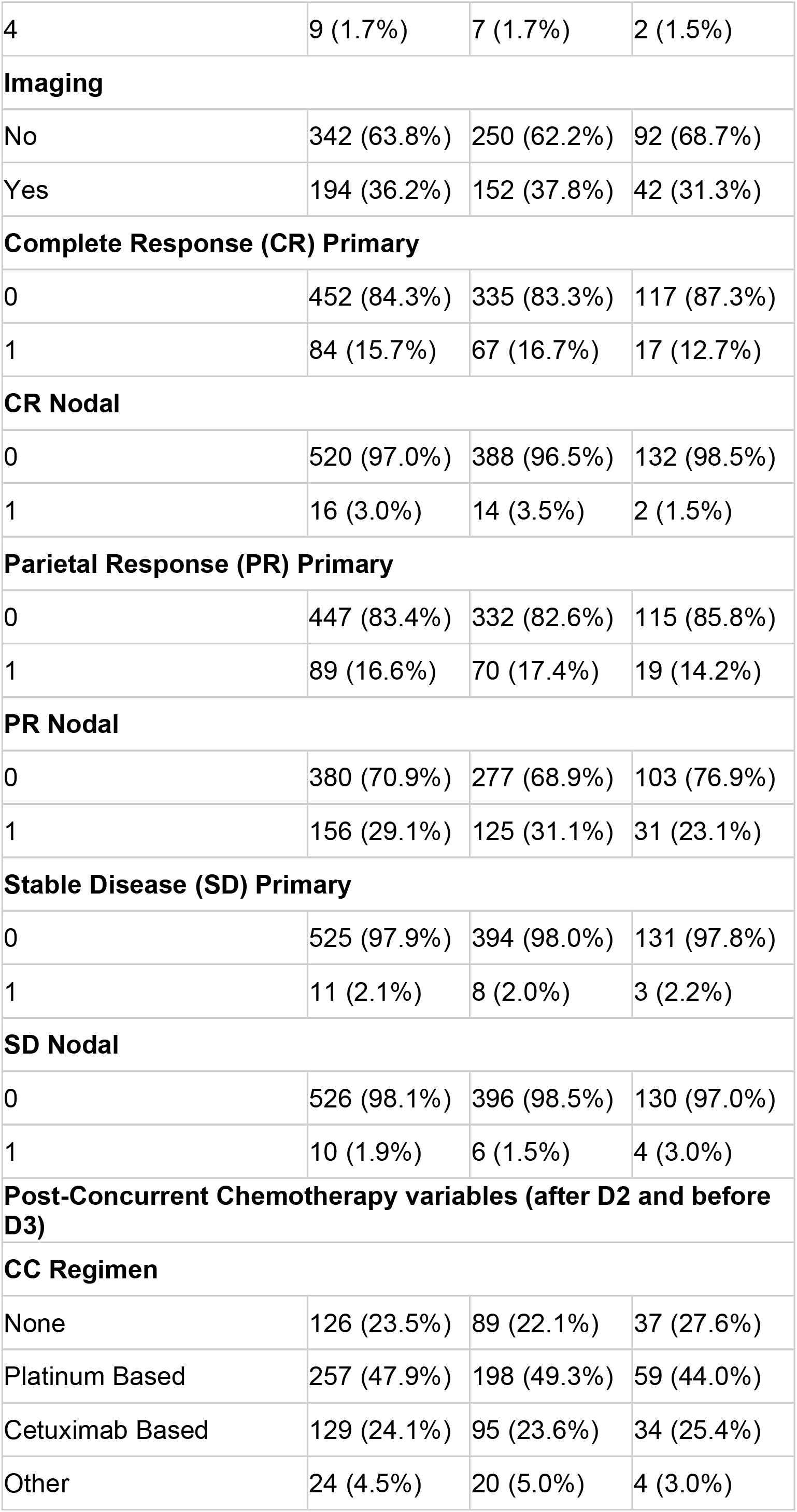
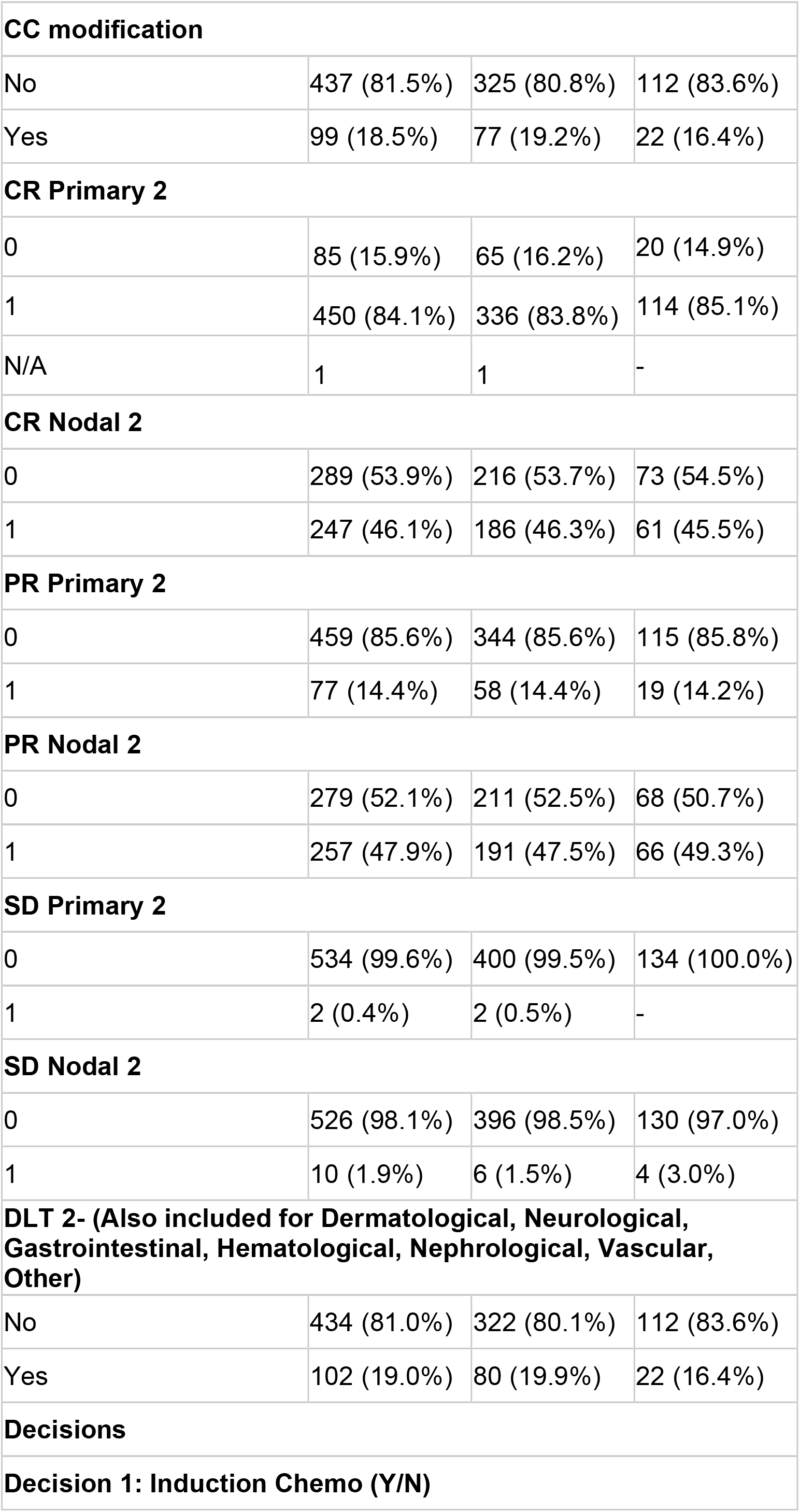
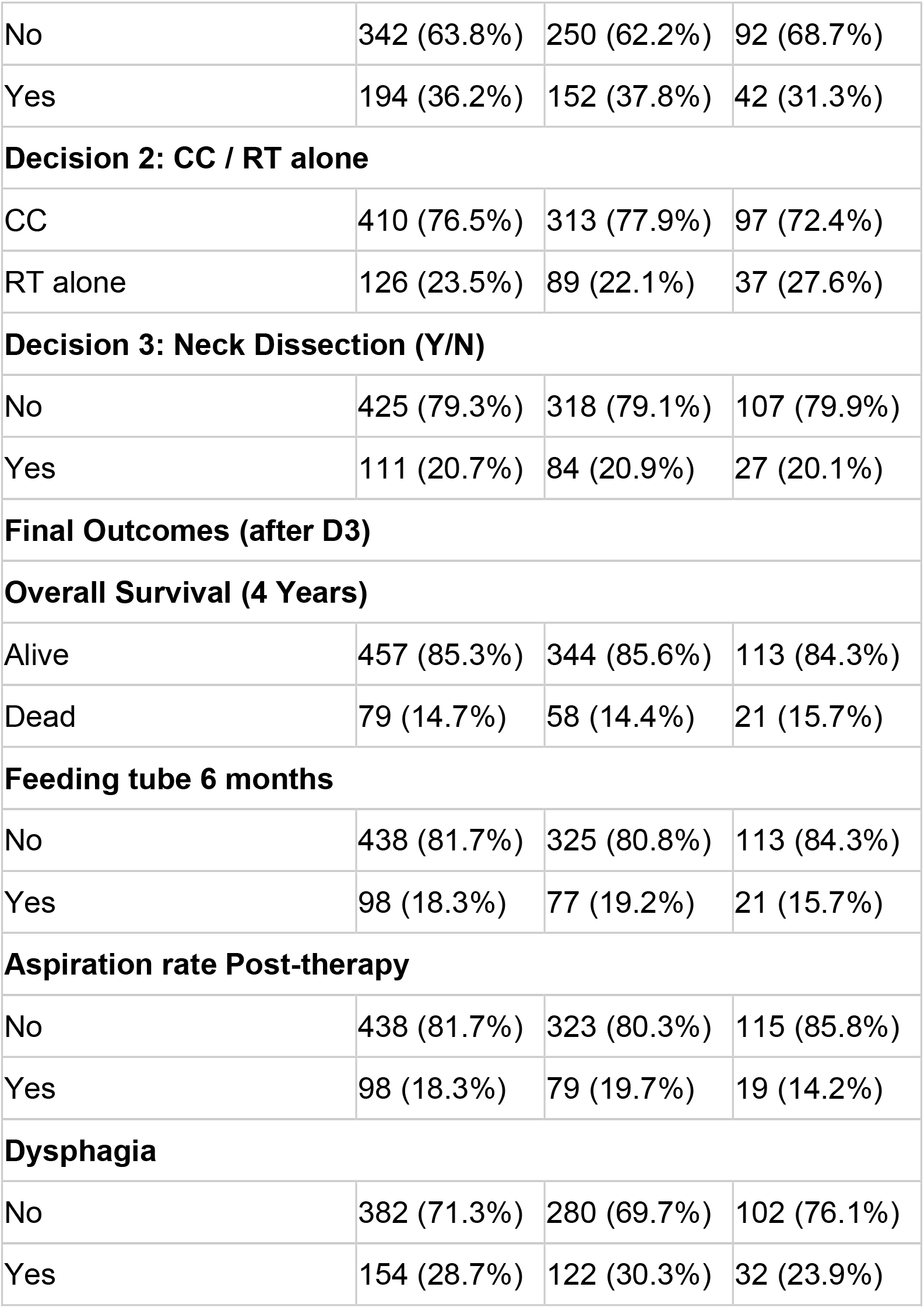
Main feature demographics for each treatment junction.

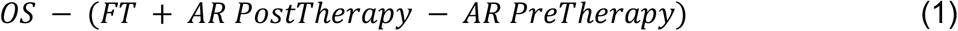

and it is used as the total reward in training the DQL models. For each of these scenarios, the models have been trained with and without the inclusion of radiomics features [17].

### Preprocessing

The dataset was split (75-25% random) into training and testing sets. To reduce radiomic feature dimensionality (∼1000) [18], we applied Principal Component Analysis (PCA) and kept the 6 top components, which explain 90% of the overall feature variance.

### DQL Modeling

A separate DQL neural network model was trained for each of the three decision points D1-3 (eFigure 1.a, in Appendix). Each model was constructed recursively based on the previous model results at the subsequent decision point, or the outcome (single or combined) in the case of D3. The models are trained to optimize the total rewards (i.e. the combined outcome of Equation (1)), without any discounting factor. We construct multiple shallow-to-deep neural networks with an increasing number of layers, until the deepest model shows poor performance due to overfitting. We sampled 1000 separate training sets from the initial training data, and trained a separate model on each of these sets, thus obtaining bootstrapped models with 95% confidence intervals.

The trained models are then used to prescribe the optimal treatment (eFigure 1 bottom, in Appendix). By prescribing an optimal treatment at each treatment junction, the DQL models construct a *digital twin* of the decision process, even though their objective is not to replicate the physicians’ decisions. The trained models and the code to compute the treatment decisions are made available on GitHub[19].

### Treatment Simulator

Because the DQL goal is not to replicate clinicians’ decisions, but to find an optimal, potentially different treatment, our evaluation includes building a Treatment Simulation (TS) model which, given a patient’s history and the prescribed treatment, predicts the outcome of that treatment. The TS consists of a model for each intermediate and final outcome measure, built using a Support Vector Classifier (SVC) and tuned via 5-fold cross-validation over the training data. The prediction accuracy of the TS with 95% confidence intervals was assessed with out of bag evaluation of 1000 models trained on stratified bootstrapped samples. The full details of the SVC are provided in the eTable 4 in the Methodology Appendix. This *patient treatment digital twin* approach enables us to simulate the results of applying the Q-learning models to patients, and compare the outcomes to the outcomes resulting from the clinicians’ decisions.

We note that although many RL algorithms learn by applying the policy in an environment, we do not use the TS in the training phase, and only limit its use to evaluation. This approach, based on learning from observation data, enables us to avoid the risk of overfitting the DQL models to the simulated environment. This results in a more objective evaluation of the learned DQL models.

### Evaluation

The TS performance is evaluated by comparing the individual DQL outcomes with those yielded by the individual physicians’ prescriptions, as well as at the whole policy level. The DQL models are then evaluated by comparing the survival and dysphagia rate yielded by the DQL treatment decisions (as computed by the TS) with the outcomes observed under physician treatment on a separate test set. To facilitate interpretation, we compute the similarity between each DQL model’s decisions and the physicians’ decisions, considering each decision point independently, and without inclusion of the treatment simulator (TS). To further support interpretation, the policy followed by each model is analyzed by computing the increase (or decrease) in prescription rate for each treatment decision compared to the physicians’ *ad-hoc* prescriptions, to express whether the model is more or less likely to prescribe a certain treatment when compared to actual physicians.

We also evaluate the DQL treatment decisions by examining compliance with the NCCN guidelines of acceptable care [20] which state that eligible advanced-stage patients (T3-4 or N1-2-3) must be prescribed chemotherapy, either induction (D1) or concurrent (D2). These guidelines/restrictions were not explicitly introduced during model training. The eMethods in the Appendix provide further DQN and TS method and model details.

We first report the performance of the treatment simulator TS, simulation performance of DQL models, and compare the DQL recommendations to the physician decision process, both in terms of overall similarity and prescription rate of each treatment decision. For reporting compliance, and to ensure quality and facilitate reproducibility, we provide a formal presentation of the TRIPOD checklist, formalized in eTable 5 in the Appendix.

## Results

The complete prediction accuracy of each TS model (with 95% confidence intervals) can be found in Table 2. The average bootstrapped prediction accuracy of the individual TS models was 87.35%, and median accuracy was 92.07%. At the whole policy level, where the TS predicts the final outcome of the complete physicians’ treatment sequence, the average prediction accuracy on the test set outcomes is 83.21%, with 83.96% accuracy for overall survival OS, and 82.46% for dysphagia DP (88.43% for feeding tube FT, and 83.58% for aspiration rate AR).

**Table 2:**
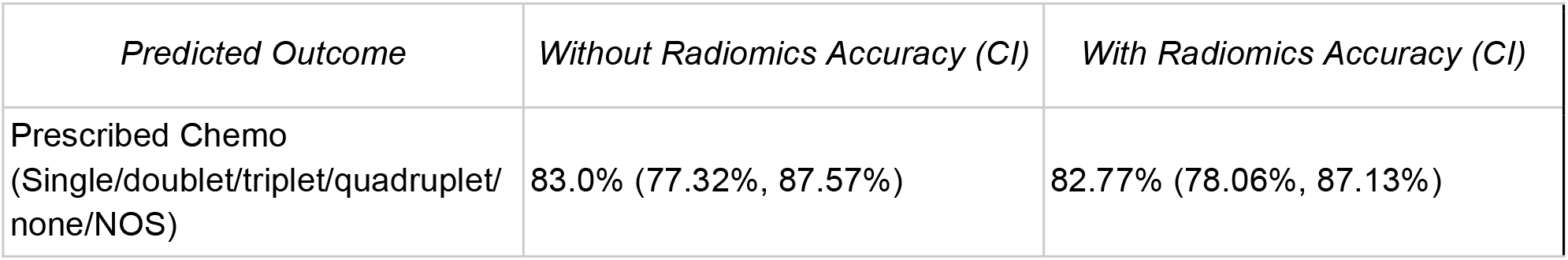
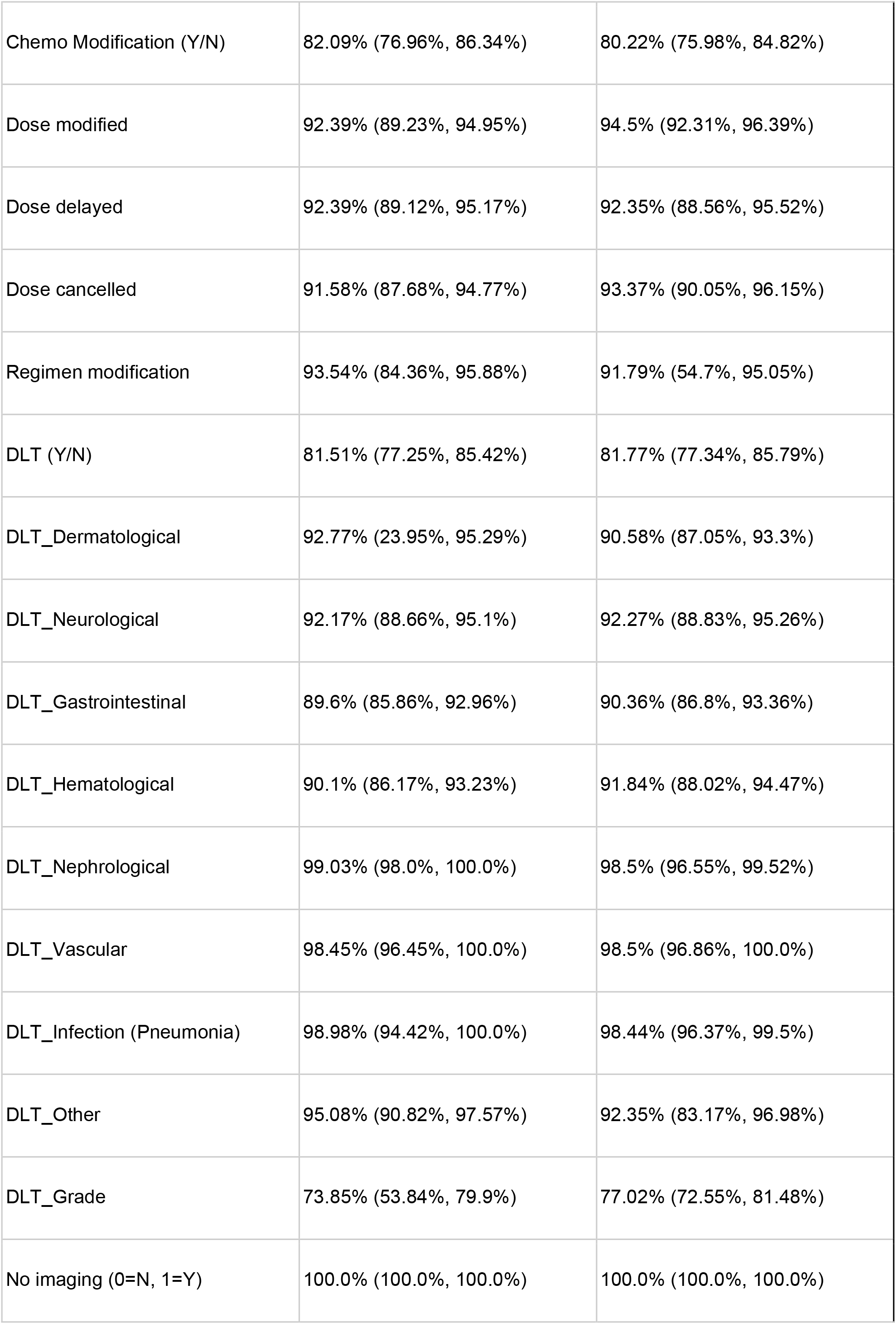
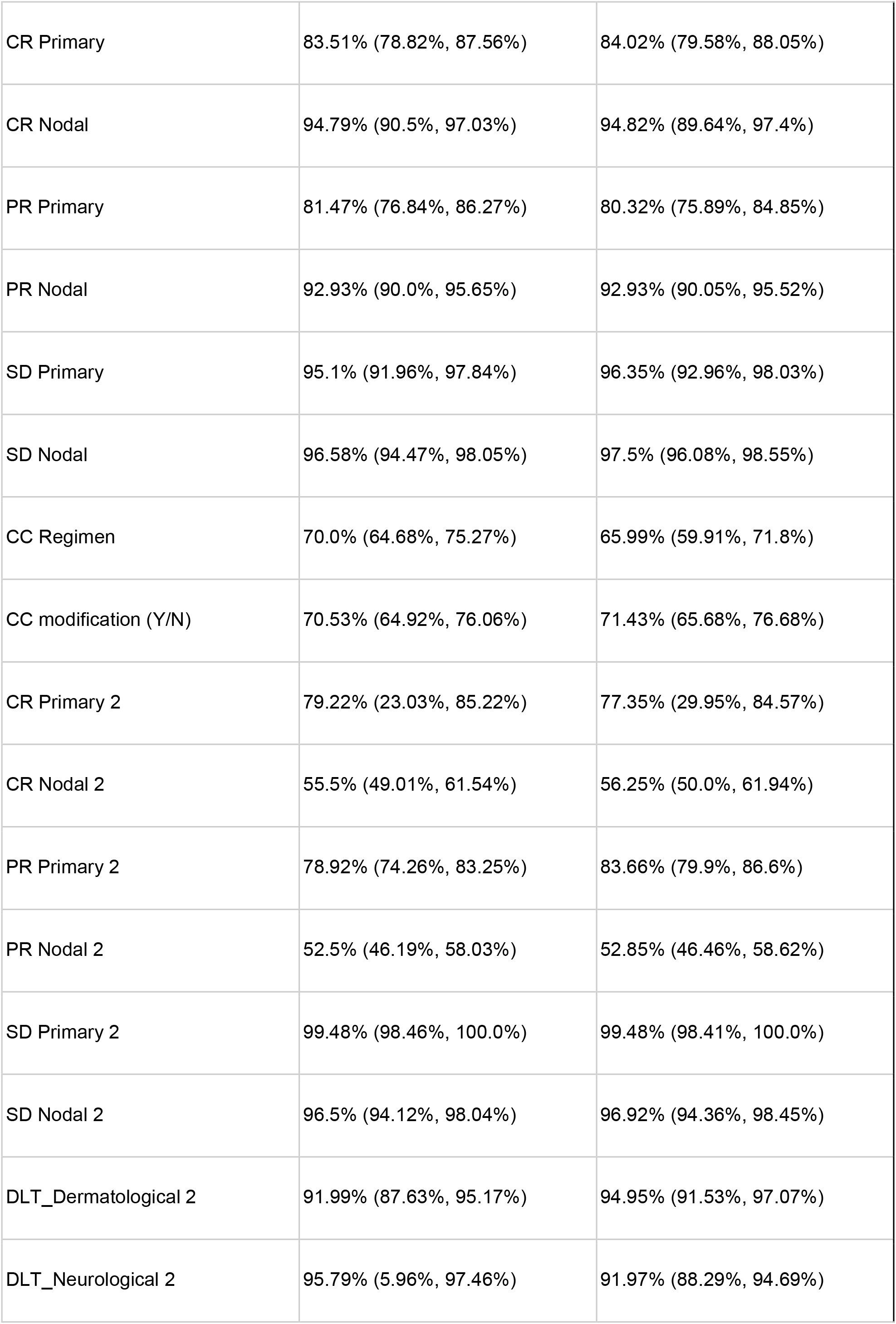
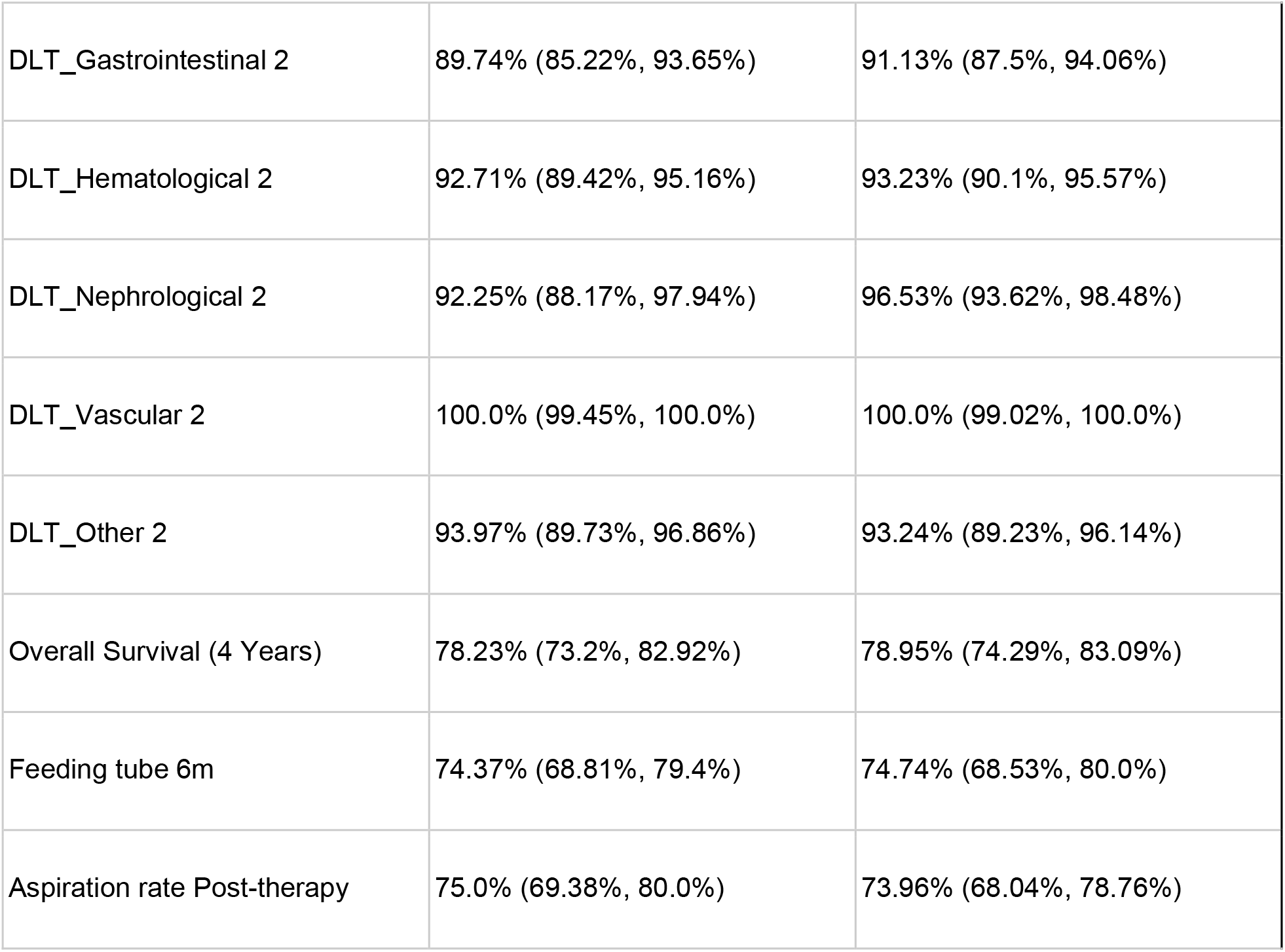
Prediction accuracy (with Confidence Intervals) result of out of bag evaluation of 1000 stratified bootstrapped samples. 95% Confidence Intervals are shown in brackets.

| *Predicted Outcome* | *Without Radiomics Accuracy (CI)* | *With Radiomics Accuracy (CI)* |
| --- | --- | --- |
| Prescribed Chemo<br>(Single/double/triplet/quadruplet/<br>none/NOS) | 83.0% (77.32%, 87.57%) | 82.77% (78.06%, 87.13%) |
| Chemo Modification (Y/N) | 82.09% (76.96%, 86.34%) | 80.22% (75.98%, 84.82%) |
| Dose modified | 92.39% (89.23%, 94.95%) | 94.5% (92.31%, 96.39%) |
| Dose delayed | 92.39% (89.12%, 95.17%) | 92.35% (88.56%, 95.52%) |
| Dose cancelled | 91.58% (87.68%, 94.77%) | 93.37% (90.05%, 96.15%) |
| Regimen modification | 93.54% (84.36%, 95.88%) | 91.79% (54.7%, 95.05%) |
| DLT (Y/N) | 81.51% (77.25%, 85.42%) | 81.77% (77.34%, 85.79%) |
| DLT_Dermatological | 92.77% (23.95%, 95.29%) | 90.58% (87.05%, 93.3%) |
| DLT_Neurological | 92.17% (88.66%, 95.1%) | 92.27% (88.83%, 95.26%) |
| DLT_Gastrointestinal | 89.6% (85.86%, 92.96%) | 90.36% (86.8%, 93.36%) |
| DLT_Hematological | 90.1% (86.17%, 93.23%) | 91.84% (88.02%, 94.47%) |
| DLT_Nephrological | 99.03% (98.0%, 100.0%) | 98.5% (96.55%, 99.52%) |
| DLT_Vascular | 98.45% (96.45%, 100.0%) | 98.5% (96.86%, 100.0%) |
| DLT_Infection (Pneumonia) | 98.98% (94.42%, 100.0%) | 98.44% (96.37%, 99.5%) |
| DLT_Other | 95.08% (90.82%, 97.57%) | 92.35% (83.17%, 96.98%) |
| DLT_Grade | 73.85% (53.84%, 79.9%) | 77.02% (72.55%, 81.48%) |
| No imaging (0=N, 1=Y) | 100.0% (100.0%, 100.0%) | 100.0% (100.0%, 100.0%) |
| CR Primary | 83.51% (78.82%, 87.56%) | 84.02% (79.58%, 88.05%) |
| CR Nodal | 94.79% (90.5%, 97.03%) | 94.82% (89.64%, 97.4%) |
| PR Primary | 81.47% (76.84%, 86.27%) | 80.32% (75.89%, 84.85%) |
| PR Nodal | 92.93% (90.0%, 95.65%) | 92.93% (90.05%, 95.52%) |
| SD Primary | 95.1% (91.96%, 97.84%) | 96.35% (92.96%, 98.03%) |
| SD Nodal | 96.58% (94.47%, 98.05%) | 97.5% (96.08%, 98.55%) |
| CC Regimen | 70.0% (64.68%, 75.27%) | 65.99% (59.91%, 71.8%) |
| CC modification (Y/N) | 70.53% (64.92%, 76.06%) | 71.43% (65.68%, 76.68%) |
| CR Primary 2 | 79.22% (23.03%, 85.22%) | 77.35% (29.95%, 84.57%) |
| CR Nodal 2 | 55.5% (49.01%, 61.54%) | 56.25% (50.0%, 61.94%) |
| PR Primary 2 | 78.92% (74.26%, 83.25%) | 83.66% (79.9%, 86.6%) |
| PR Nodal 2 | 52.5% (46.19%, 58.03%) | 52.85% (46.46%, 58.62%) |
| SD Primary 2 | 99.48% (98.46%, 100.0%) | 99.48% (98.41%, 100.0%) |
| SD Nodal 2 | 96.5% (94.12%, 98.04%) | 96.92% (94.36%, 98.45%) |
| DLT_Dermatological 2 | 91.99% (87.63%, 95.17%) | 94.95% (91.53%, 97.07%) |
| DLT_Neurological 2 | 95.79% (5.96%, 97.46%) | 91.97% (88.29%, 94.69%) |
| DLT_Gastrointestinal 2 | 89.74% (85.22%, 93.65%) | 91.13% (87.5%, 94.06%) |
| DLT_Hematological 2 | 92.71% (89.42%, 95.16%) | 93.23% (90.1%, 95.57%) |
| DLT_Nephrological 2 | 92.25% (88.17%, 97.94%) | 96.53% (93.62%, 98.48%) |
| DLT_Vascular 2 | 100.0% (99.45%, 100.0%) | 100.0% (99.02%, 100.0%) |
| DLT_Other 2 | 93.97% (89.73%, 96.86%) | 93.24% (89.23%, 96.14%) |
| Overall Survival (4 Years) | 78.23% (73.2%, 82.92%) | 78.95% (74.29%, 83.09%) |
| Feeding tube 6m | 74.37% (68.81%, 79.4%) | 74.74% (68.53%, 80.0%) |
| Aspiration rate Post-therapy | 75.0% (69.38%, 80.0%) | 73.96% (68.04%, 78.76%) |

The baseline outcomes observed under physician care are 85.57% (train set) and 84.33% (test set) OS rate, and 69.65% (train) and 76.12% (test) (absence of) DP rate.

The complete simulation performance (with 95% confidence intervals) on training and test set of the DQL models is presented in eTable 2 in the Appendix. Our best-performing DQL model is the 2-layer Neural Network (NN), which considers the combined survival and toxicity outcomes (OS+DP), without radiomics features. This model yielded a median OS improvement, compared to the baseline model, of +2.74% (95% CI: [-0.25%, +6.47%]) (train) and +3.73% (95% CI: [-0.75%, +8.96%]) (test), with an absolute highest survival rate of 88.31% (CI: [85.32%, 92.04%]) (train) and 88.06% (CI: [83.58%, 93.53%]) (test). With respect to Dysphagia (DP), the same 2-layer model showed a +3.98% (CI: [-1.24%, +9.2%]) improvement on training data, with 73.63% (CI: [68.41%, 78.86%]) of simulated patients not exhibiting DP under the model’s treatment decisions. This 2-layer model yielded a +0.75% (CI: [-4.48%, +6.72%]) improvement on the test set, from the baseline 76.12% to 76.87% (CI: [71.64%, 82.84%]).

The 2-layer model provided the best lower bounds on performance on all outcomes, on both training and test data, while still providing good median performance, and was therefore considered the best-performing one.

In order to assess model parsimony (i.e., the minimum number of layers that still maintains equivalent predictive performance), Figure 2 shows a comparison of DQL models with different number of layers, on simulated test data for the combined outcome models (OS and DP), and with and without radiomics features. Broadly, neither simpler models with <2 layers nor models with >4 layers performed as effectively as the 2 layer model.

**Figure 2.**
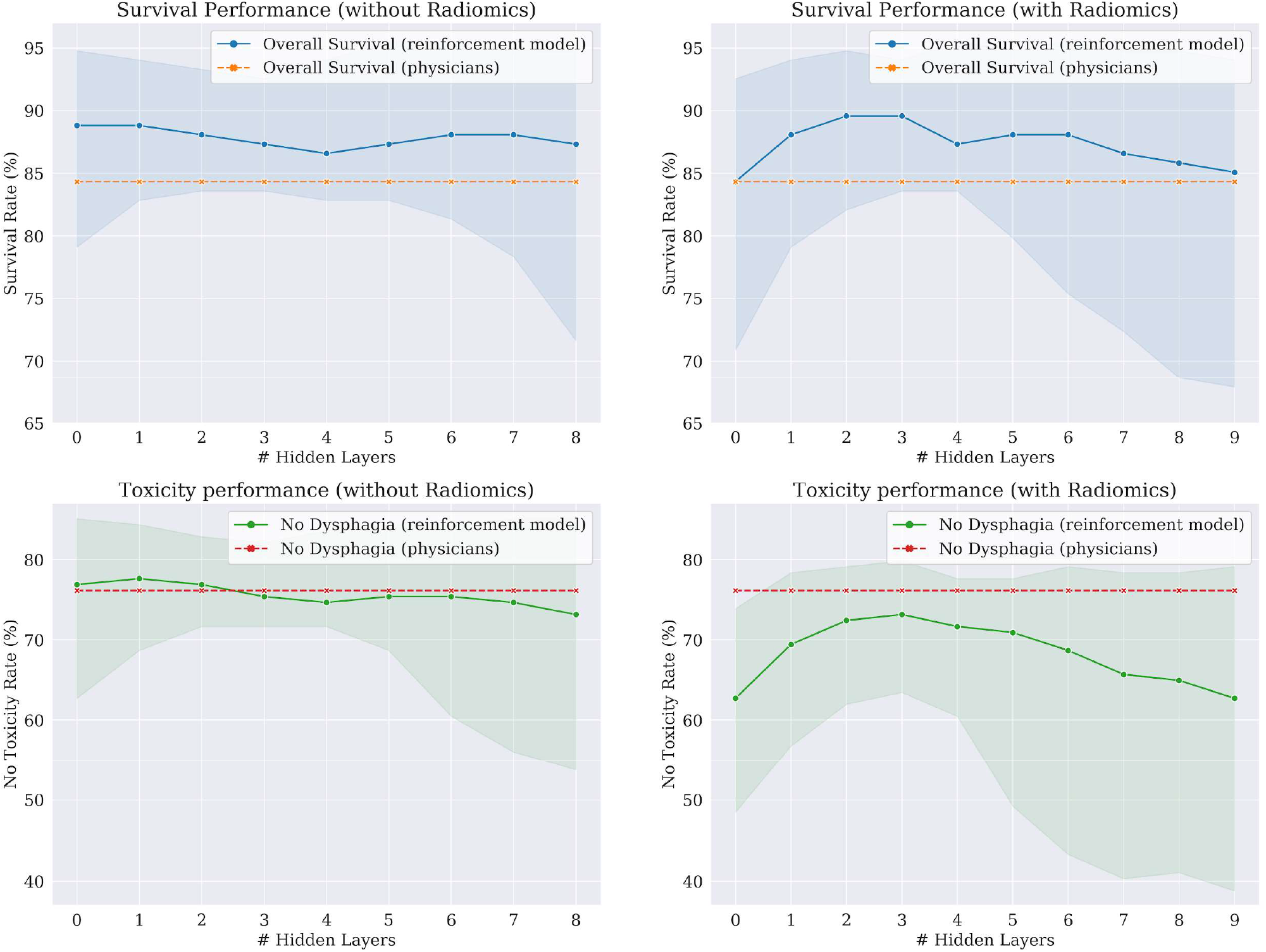
Model performance for the combined outcome (OS+DP) models without (left) and with radiomics (right) The figure shows performance for OS (overall survival, top) and Dysphagia (bottom) with varying number of layers showing treatment simulation results on test data. Continuous lines show average performance of the bootstrapped models, while highlighted areas represent the 95% confidence intervals. Dashed lines show the empirical patient outcomes observed under the physicians’ decisions. Note that models with 0 layers (linear models) are insufficient to capture the decision process, whereas models with more than 8 to 9 layers are overfitting the data. Furthermore, models without radiomics show better simulated outcomes and lower variance than models with radiomics.

The overall similarity rates (with 95% CI) with respect to physicians’ treatment “twinning”, both per decision and overall, for all models, are presented in eTable 3 in the Appendix. In terms of overall similarity of the *in silico* Q-learning treatment policies to those actually delivered *ad hoc in vivo* by physicians, our best model (OS+DP, 2-layers, no radiomics) showed an overall 70.4% (95% CI: [65.34%, 73.63%]) similarity to the physician decisions on the training set (i.e. the model chooses the same treatment as the physicians for 70.4% of the considered treatment decisions), and 69.65% (CI: [63.43%, 73.38%]) on the test set, although another model (the 3-layer OS+DP model without radiomics, which performed consistently worse in simulation for all outcomes) did show higher similarity rates.

The demographics (with 95% CI) of T- and N-stage of patients in the test set separated by chemotherapeutic treatment prescribed by the best-performing model are presented in Table 3.

**Table 3:**
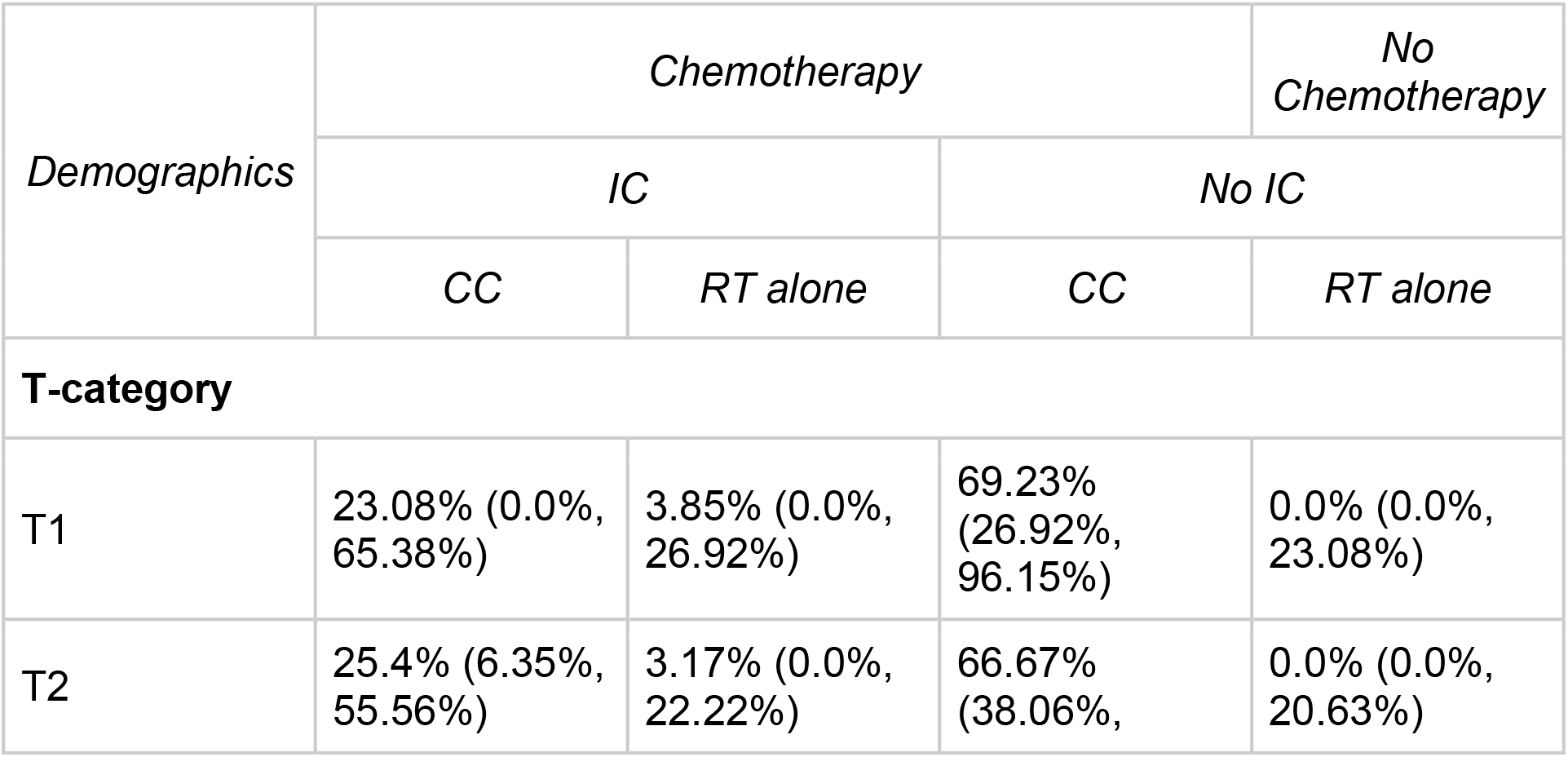
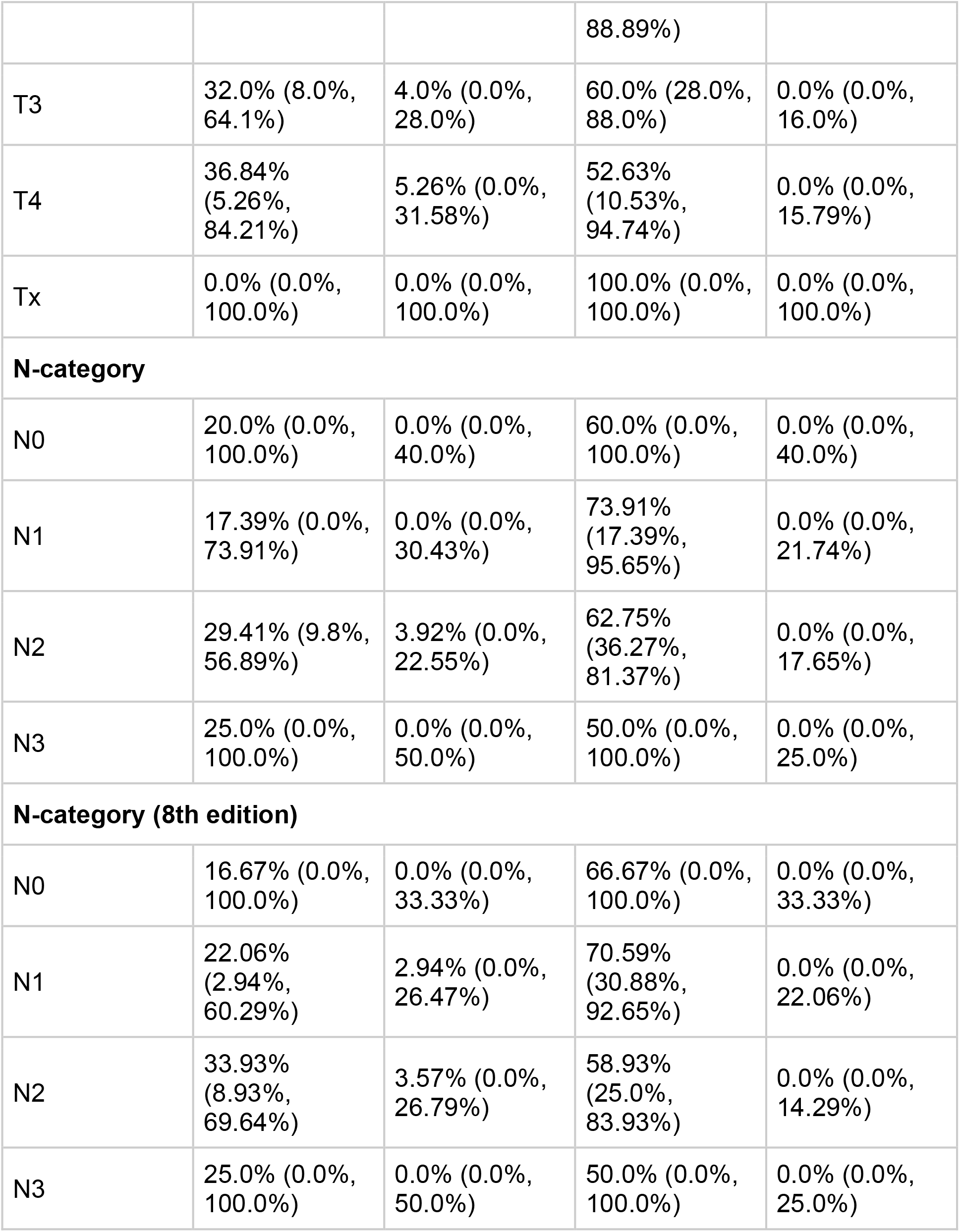
Tumor stage demographics of patients based on the chemotherapeutic treatment decisions of the best-performing model.

The rate at which models choose a certain policy with respect to the physicians’ treatment rate at each decision point is shown in Figure 3. The best-performing model in terms of simulated outcomes (OS+DP, 2 layers, no radiomics) had a higher induction chemotherapy (IC, D1) rate (2.99% increase in prescription rate compared to physicians’ prescriptions, for a total of 46 patients; 95% CI: [-14.93%, +26.88%]), and one of the highest concurrent chemo (CC, D2) rates (21.64% increase, 126 patients; CI: [-2.99%, 27.61%]), and the lowest neck dissection (ND, D3) rate (20.15%% decrease, 0 patients; CI: [-20.15%, -11.19%]).

**Figure 3.**
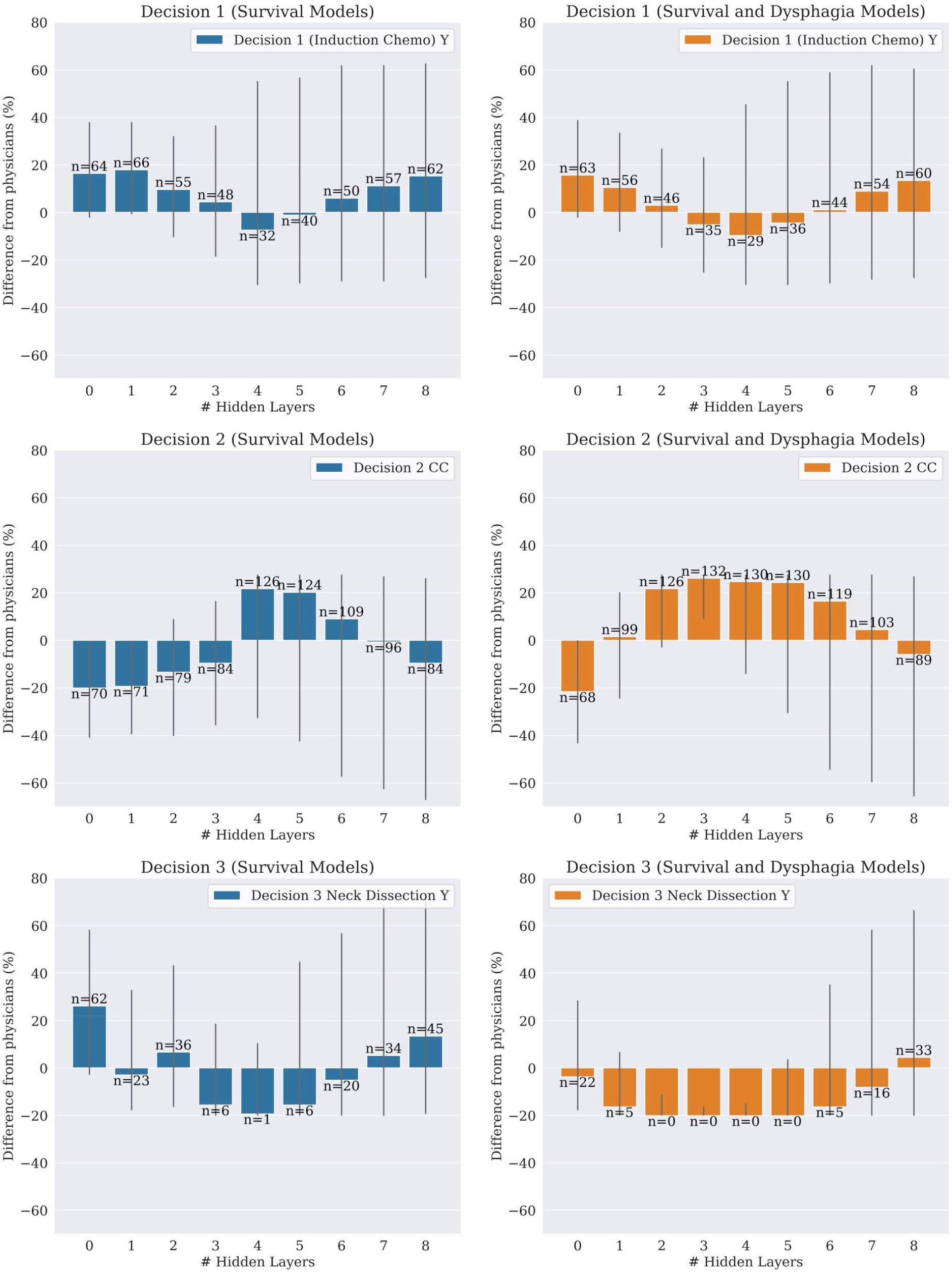
Absolute increase (or decrease) of treatment decision rate compared to physicians’ decisions. The plots refer to decisions 1 (top), 2 (middle), and 3 (bottom), on the test set, and for models considering only OS as an outcome measure (left) or OS+DP (right) without radiomics. Grey lines represent the 95% confidence intervals. The numbers on top of the bars show for how many patients (out of 134) the Q-learning model recommended that treatment. The IC prescription rate varies in a similar way between OS and OS+DP, CC is significantly more frequent in OS+DP models, while ND is consistently less frequent in OS+DP.

The training time for a single DQL model did not significantly vary between shallower and deeper NNs, and was just a few minutes on average for a complete model. With 1000 sample bootstrapping, the training time was accordingly longer; however, these models are computationally inexpensive, and can be deployed virtually in real-time.

## Discussion

The high average, median, and overall accuracy provided by the Treatment Simulator (TS) in predicting the outcomes of treatments indicate that the TS is a valid digital twin for the treatment process when predicting the outcome of a treatment sequence. Our results also indicate that the Q-learning models indeed capture the nature of the dynamic treatment problem and provide a valid solution. Our models showed consistent improvements for all the outcome variables taken into account, as well as moderate similarity to physicians’ decisions. Overall, these results indicate that DQL modeling can serve as a digital twin of the treatment decision process, and TS modeling can serve as a digital twin of the patient treatment. When combined, DQL and TS constitute a valid patient-physician *digital twin dyad* for optimal policy determination in sequential systemic and locoregional therapy of oropharyngeal squamous carcinomas.

Furthermore, our results show that the DQL models that consider OS+DP outperform models considering only OS in terms of simulated survival rate. Since the absence of Dysphagia (FT or AR) symptoms is positively correlated with survival, maximizing these indirectly helps maximize OS-model performance as well.

Moreover, OS+DP models also show higher similarity to actual physician decisions, as they represent a more granular approximation of the decision process than models that include only OS as an outcome, including more of the variables considered by the physician when choosing an optimal treatment.

Surprisingly, given the abundance of data on radiomics models for head and neck cancer [21, 22, 23, 24, 25, 26, 27, 28, 29], Q-learning models *without* radiomics yielded a better performance than models that included radiomics, in terms of simulated outcomes and variance, for both training and testing data. The most evident example is given by the simulated Dysphagia: none of the models trained with radiomics features managed to improve the outcome observed following physicians’ treatment in the test set (Figure 2, bottom-right). From these results, addition of textural features *failed* to improve model performance, and instead significantly increased the performance variance of the bootstrapped models.

Our findings also justify the choice of a Deep NN model instead of a regular linear model: whereas by using DQL we reduce model parsimony, we can see that the results of the linear models (i.e. the 0-hidden-layers NNs) are comparatively suboptimal to deeper models, both in terms of simulated performance, confidence intervals variance, and similarity.

Furthermore, per Table 3, the best performing model did not violate the standard of care regarding chemotherapeutic treatment of patients, since all patients with T3-4 or N1-2-3 stage cancer were prescribed either induction (D1) or concurrent (D2) chemotherapy, with the majority being prescribed at least concurrent chemotherapy, showing clinical applicability was maintained.

When comparing OS-only models versus OS+DP models, the prescription rates presented in Figure 3 showed two separate trends for the two categories: whereas the induction chemo (IC, D1) prescription rate varied similarly for OS and OS+DP models, the rates of concurrency chemo (CC, D2) and neck dissection (ND, D3) were significantly different between the two categories. In general, OS+DP models tended to have a higher rate of CC (D2) and a lower rate of ND (D3). In other words, *models that also consider toxicity as an outcome measure balanced a more aggressive chemotherapeutic treatment with a lower rate of neck dissection*, which is consistent with the known positive correlation between surgery and dysphagia symptoms such as feeding tube and aspiration rate.

## Limitations

While the proposed approach was shown to be effective in dynamically selecting optimal treatment strategy for OPC patients, it is not without limitations. Due to the retrospective nature of the dataset, our Q-learning models had to be evaluated through the Treatment Simulator, a Supervised Learning model, which might be seen as self-referential. The TS is, however, a necessary approach before prospective application, because evaluating the models based on physician similarity alone would not reflect the purpose of DQL. Intuitively, the goal of DQL is not to replicate the decisions taken by physicians in the dataset, but to learn from these decisions and their effect in order to discern between optimal and non-optimal choices with respect to a given outcome measure.

Furthermore, because we train our *digital twin dyad* on a representative cohort from a single cancer center (M.D. Anderson Cancer Center), the physician decision/prescribing heuristics reflected may not be fully generalizable to other facilities with other practitioners. The physician prescriptions at the center are, however, aligned with the state of the art in the field. In particular, we note that whereas two studies [30, 31] have questioned the relevance of induction chemotherapy to treatment, both studies have failed to accrue and thus are null. Our modeling approach could conceivably be implemented and extended to generate similar *digital twin dyad*s across other or multiple institutions.

## Conclusions

In conclusion, we constructed a deep Q-learning modeling approach to make optimized sequential treatment decisions based on a set of desired outcomes in head and neck cancer therapy, and paired it with a simulation of the treatment process for evaluation purposes. This modeling approach represents, to our knowledge, the first application of deep-Q-learning with simulation as a *digital twin dyad* to simultaneously represent both s*tate-specific patient data* and *physician/prescriber policies* for head and neck squamous carcinoma. Furthermore, this work is the first reported implementation of deep-Q-learning for dysphagia and survival composite-outcome modeling. Our approach further demonstrates the technical feasibility of such a *digital twin dyad*, and provides a benchmarking dataset and relevant code for model dissemination, site-specific implementation, and iterative model improvement. Carrying out a prospective clinical study application could further confirm the validity of this approach as part of the standard of care.

## Data Availability

The dataset used in the data analysis is publicly available.

https://doi.org/10.6084/m9.figshare.13235453.v2

## Acknowledgements

We thank all members of the Electronic Visualization Laboratory, members of the MD Anderson Head and Neck Cancer Quantitative Imaging Collaborative Group, and our collaborators at the University of Iowa and University of Minnesota.

## †-Co-author specific contributions

All listed co-authors performed the following:

1. *Substantial contributions to the conception or design of the work; or the acquisition, analysis, or interpretation of data for the work;*
2. *Drafting the work or revising it critically for important intellectual content;*
3. *Final approval of the version to be published;*
4. *Agreement to be accountable for all aspects of the work in ensuring that questions related* to the accuracy or integrity of any part of the work are appropriately investigated and resolved.

Specific additional individual cooperative effort contributions to study/manuscript design/execution/interpretation, in addition to all criteria above are listed as follows:

- *ET, XZ, GC, DF, AW, GEM - designed and developed the machine learning models, data extraction and curation, statistical analysis, and interpretation*
- *LVD, ASRM, DF - direct patient care provision, direct clinical data collection; interpretation and analytic support*
- *GC - supervised statistical analysis, data extraction, analytic support, guarantor of statistical quality*
- *ET, XZ, AW, GC, LVD, ASRM, CDF, GEM - manuscript writing and editing*
- *XZ, GC, CDF, GEM - primary investigator(s); conceived, coordinated, and directed all study activities, responsible for data collection, project integrity, manuscript content and editorial oversight and correspondence*

## Data Sharing Statement

The dataset used in the data analysis is publicly available [32].

