## Appendix for "Optimal policy determination in sequential systemic and locoregional therapy of oropharyngeal squamous carcinomas: A patient-physician digital twin dyad with deep Q-learning for treatment selection"

**Appendix: Table of contents**

**eTable 1: Feature demographics for each treatment junction.** 2

**eTable 2: Absolute improvement over physicians' results with treatment simulation, per model, without and with radiomics, and for different model outcomes (OS vs. OS+DP).** 12

**eTable 3: Model similarity to physicians' decisions on training and testing data, without radiomics and with radiomics, and for different model outcomes (OS and OS+DP).** 16

**eMethodology** 22

**eFigure 1. Overview of DQL model training and usage.** 27

**etable 4: SVC details** 29

**eTable 5: TRIPOD checklist** 34

**eReferences** 39

**eTable 1: Feature demographics for each treatment junction.**

Demographics are shown for all 536 patients as well as for training and test set separately.

| *Characteristic* | *All Patients (536)* | *Training set (402)* | *Testing set (134)* |
|---|---|---|---|
| **Pre-treatment variables (before D1)** | | | |
| **Age at Diagnosis (Calculated)** | | | |
| Mean (SD) | 58.9 (9.5%) | 58.5 (9.4%) | 60.2 (9.6%) |
| **Pathological Grade** | | | |
| I | 6 (1.4%) | 2 (0.6%) | 4 (3.5%) |
| II | 154 (35.2%) | 114 (35.3%) | 40 (35.1%) |
| III | 274 (62.7%) | 206 (63.8%) | 88 (59.6%) |
| IV | 3 (0.7%) | 1 (0.3%) | 2 (1.8%) |
| N/A | 99 | 79 | 20 |
| **Gender** | | | |
| Female | 65 (12.1%) | 47 (11.7%) | 18 (13.4%) |
| Male | 471 (87.9%) | 355 (88.3%) | 116 (86.6%) |
| **HPV/P16 status** | | | |
| Negative | 43 (8%) | 33 (8.2%) | 10 (7.5%) |
| Positive | 305 (56.9%) | 228 (56.7%) | 77 (57.5%) |
| Unknown | 188 (35.1%) | 141 (35.1%) | 47 (35.1%) |
| **T-category** | | | |
| T1 | 113 (21.1%) | 87 (21.6%) | 26 (19.4%) |
| T2 | 219 (40.9%) | 156 (38.8%) | 63 (47.0%) |
| T3 | 116 (21.6%) | 91 (22.6%) | 25 (18.7%) |

| | | | |
|---|---|---|---|
| T4 | 86 (16.0%) | 67 (16.7%) | 19 (14.2%) |
| Tis | - | - | - |
| Tx | 2 (0.4%) | 1 (0.2%) | 1 (0.7%) |
| **N-category** | | | |
| N0 | 19 (3.5%) | 14 (3.5%) | 5 (3.7%) |
| N1 | 62 (11.6%) | 39 (9.7%) | 23 (17.2%) |
| N2 | 438 (81.7%) | 336 (83.6%) | 102 (76.1%) |
| N3 | 17 (3.2%) | 13 (3.2%) | 4 (3.0%) |
| **N-category (8th edition)** | | | |
| N0 | 20 (3.7%) | 14 (3.5%) | 6 (4.5%) |
| N1 | 249 (46.5%) | 181 (45%) | 68 (50.7%) |
| N2 | 250 (46.6%) | 194 (48.3%) | 56 (41.8%) |
| N3 | 17 (3.2%) | 13 (3.2%) | 4 (3.0%) |
| **AJCC 7th edition** | | | |
| II | 9 (1.7%) | 6 (1.5%) | 3 (2.2%) |
| III | 64 (11.9%) | 39 (9.7%) | 25 (18.7%) |
| IV | 463 (86.4%) | 357 (88.8%) | 106 (79.1%) |
| **AJCC 8th edition** | | | |
| I | 186 (34.8%) | 137 (34.2%) | 49 (36.8%) |
| II | 81 (15.2%) | 63 (15.7%) | 18 (13.5%) |
| III | 64 (12.0%) | 44 (11.0%) | 20 (15.0%) |
| IV | 203 (38.0%) | 157 (39.2%) | 46 (34.6%) |
| N/A | 2 | 1 | 1 |
| **Smoking status at Diagnosis** | | | |
| Current | 115 (21.5%) | 85 (21.1%) | 30 (22.4%) |

| | | | |
|---|---|---|---|
| Former | 203 (37.9%) | 151 (37.6%) | 52 (38.8%) |
| Never | 218 (40.7%) | 166 (41.3%) | 52 (38.8%) |
| **Smoking status (Packs/Year)** | | | |
| Mean (SD) | 17.7 (23.7) | 16.7 (22.9) | 20.5 (26.0) |
| N/A | 28 | 21 | 7 |
| **Aspiration rate Pre-therapy** | | | |
| No | 520 (97.0%) | 388 (96.5%) | 132 (98.5%) |
| Yes | 16 (3.0%) | 14 (3.5%) | 2 (1.5%) |
| **Number of Affected Lymph nodes** | | | |
| Mean (SD) | 2.0 (1.3) | 2.1 (1.3) | 1.8 (1.0) |
| **Tumor Laterality** | | | |
| Bilateral | 21 (3.9%) | 16 (4.0%) | 5 (3.7%) |
| Left | 242 (45.1%) | 188 (46.8%) | 54 (40.3%) |
| Right | 273 (50.9%) | 198 (49.3%) | 75 (56.0%) |
| **Tumor subsite** | | | |
| Base Of Tongue | 266 (49.6%) | 204 (50.7%) | 62 (46.3%) |
| GlossoPharyngeal Sulcus | 10 (1.9%) | 10 (2.5%) | - |
| Not Otherwise Specified | 31 (5.8%) | 24 (6.0%) | 7 (5.2%) |
| Soft Palate | 6 (1.1%) | 6 (1.5%) | - |
| Tonsil | 223 (41.6%) | 158 (39.3%) | 65 (48.5%) |
| **Race** | | | |
| African American / Black | 16 (3.0%) | 10 (2.5%) | 6 (4.5%) |
| Asian | 4 (0.7%) | 3 (0.7%) | 1 (0.7%) |
| Hispanic / Latino | 21 (3.9%) | 17 (4.2%) | 4 (3.0%) |
| Not Otherwise Specified | 5 (0.9%) | 3 (0.7%) | 2 (1.5%) |

| | | | |
|---|---|---|---|
| Native American | 1 (0.2%) | 1 (0.2%) | - |
| White / Caucasian | 489 (91.2%) | 368 (91.5%) | 121 (90.3%) |
| **Post-Induction Therapy variables (after D1 and before D2)** | | | |
| **Prescribed Chemo** | | | |
| None | 342 (63.8%) | 250 (62.2%) | 92 (68.7%) |
| Doublet | 41 (7.6%) | 32 (8.0%) | 9 (6.7%) |
| Triplet | 143 (26.7%) | 111 (27.6%) | 32 (23.9%) |
| Quadruplet | 7 (1.3%) | 7 (1.7%) | - |
| Not Otherwise Specified | 3 (0.6%) | 2 (0.5%) | 1 (0.7%) |
| **Chemo Modification** | | | |
| Yes | 85 (15.9%) | 65 (16.2%) | 20 (14.9%) |
| No | 451 (84.1%) | 337 (83.8%) | 114 (85.1%) |
| **Modification Type** | | | |
| No Dose Adjustment | 451 (84.1%) | 336 (83.6%) | 115 (85.8%) |
| Dose Modified | 21 (3.9%) | 16 (4.0%) | 5 (3.7%) |
| Dose Delayed | 10 (1.9%) | 9 (2.2%) | 1 (0.7%) |
| Dose Cancelled | 18 (3.4%) | 13 (3.2%) | 5 (3.7%) |
| Dose Delayed & Modified | 6 (1.1%) | 5 (1.2%) | 1 (0.7%) |
| Regimen Modification | 29 (5.4%) | 22 (5.5%) | 7 (5.2%) |
| Unknown | 1 (0.2%) | 1 (0.2%) | - |
| **Dose Limiting Toxicity** | | | |
| No | 441 (82.3%) | 329 (81.8%) | 112 (83.6%) |
| Yes | 95 (17.7%) | 73 (18.2%) | 22 (16.4%) |
| **DLT - Dermatological** | | | |
| 0 | 506 (94.4%) | 383 (95.3%) | 123 (91.8%) |

| | | | |
|---|---|---|---|
| 1 | 24 (4.5%) | 15 (3.7%) | 9 (6.7%) |
| 2 | 4 (0.7%) | 2 (0.5%) | 2 (1.5%) |
| 3 | 2 (0.4%) | 2 (0.5%) | - |
| **DLT - Neurological** | | | |
| 0 | 515 (96.1%) | 386 (96.0%) | 129 (96.3%) |
| 1 | 17 (3.2%) | 13 (3.2%) | 4 (3.0%) |
| 2 | 3 (0.6%) | 2 (0.5%) | 1 (0.7%) |
| 3 | 1 (0.2%) | 1 (0.2%) | - |
| **DLT- Gastrointestinal** | | | |
| 0 | 504 (94.0%) | 377 (93.8%) | 127 (94.8%) |
| 1 | 27 (5.0%) | 20 (5.0%) | 7 (5.2%) |
| 2 | 2 (0.4%) | 2 (0.5%) | - |
| 3 | 3 (0.6%) | 3 (0.7%) | - |
| **DLT - Hematological** | | | |
| 0 | 509 (95.0%) | 383 (95.3%) | 126 (94.0%) |
| 1 | 26 (4.9%) | 18 (4.5%) | 8 (6.0%) |
| 3 | - | - | - |
| 4 | 1 (0.2%) | 1 (0.2%) | - |
| **DLT - Nephrological** | | | |
| 0 | 533 (99.4%) | 399 (99.3%) | 134 (100.0%) |
| 1 | 3 (0.6%) | 3 (0.7%) | - |
| **DLT - Vascular** | | | |
| 0 | 534 (99.6%) | 401 (99.8%) | 133 (99.3%) |
| 1 | 1 (0.2%) | - | 1 (0.7%) |
| 3 | 1 (0.2%) | 1 (0.2%) | - |

| | | | |
|---|---|---|---|
| **DLT - Infection (Pneumonia)** | | | |
| 0 | 535 (99.8%) | 401 (99.8%) | 134 (100.0%) |
| 1 | 1 (0.2%) | 1 (0.2%) | - |
| **DLT - Grade** | | | |
| 0 | 446 (83.2%) | 334 (83.1%) | 112 (83.6%) |
| 1 | 7 (1.3%) | 6 (1.5%) | 1 (0.7%) |
| 2 | 33 (6.2%) | 26 (6.5%) | 7 (5.2%) |
| 3 | 41 (7.6%) | 29 (7.2%) | 12 (9.0%) |
| 4 | 9 (1.7%) | 7 (1.7%) | 2 (1.5%) |
| **Imaging** | | | |
| No | 342 (63.8%) | 250 (62.2%) | 92 (68.7%) |
| Yes | 194 (36.2%) | 152 (37.8%) | 42 (31.3%) |
| **Complete Response (CR) Primary** | | | |
| 0 | 452 (84.3%) | 335 (83.3%) | 117 (87.3%) |
| 1 | 84 (15.7%) | 67 (16.7%) | 17 (12.7%) |
| **CR Nodal** | | | |
| 0 | 520 (97.0%) | 388 (96.5%) | 132 (98.5%) |
| 1 | 16 (3.0%) | 14 (3.5%) | 2 (1.5%) |
| **Parietal Response (PR) Primary** | | | |
| 0 | 447 (83.4%) | 332 (82.6%) | 115 (85.8%) |
| 1 | 89 (16.6%) | 70 (17.4%) | 19 (14.2%) |
| **PR Nodal** | | | |
| 0 | 380 (70.9%) | 277 (68.9%) | 103 (76.9%) |
| 1 | 156 (29.1%) | 125 (31.1%) | 31 (23.1%) |
| **Stable Disease (SD) Primary** | | | |

| | | | |
|---|---|---|---|
| 0 | 525 (97.9%) | 394 (98.0%) | 131 (97.8%) |
| 1 | 11 (2.1%) | 8 (2.0%) | 3 (2.2%) |
| **SD Nodal** | | | |
| 0 | 526 (98.1%) | 396 (98.5%) | 130 (97.0%) |
| 1 | 10 (1.9%) | 6 (1.5%) | 4 (3.0%) |
| **Post-Concurrent Chemotherapy variables (after D2 and before D3)** | | | |
| **CC Regimen** | | | |
| None | 126 (23.5%) | 89 (22.1%) | 37 (27.6%) |
| Platinum Based | 257 (47.9%) | 198 (49.3%) | 59 (44.0%) |
| Cetuximab Based | 129 (24.1%) | 95 (23.6%) | 34 (25.4%) |
| Other | 24 (4.5%) | 20 (5.0%) | 4 (3.0%) |
| **CC modification** | | | |
| No | 437 (81.5%) | 325 (80.8%) | 112 (83.6%) |
| Yes | 99 (18.5%) | 77 (19.2%) | 22 (16.4%) |
| **CR Primary 2** | | | |
| 0 | 85 (15.9%) | 65 (16.2%) | 20 (14.9%) |
| 1 | 450 (84.1%) | 336 (83.8%) | 114 (85.1%) |
| N/A | 1 | 1 | - |
| **CR Nodal 2** | | | |
| 0 | 289 (53.9%) | 216 (53.7%) | 73 (54.5%) |
| 1 | 247 (46.1%) | 186 (46.3%) | 61 (45.5%) |
| N/A | - | - | - |
| **PR Primary 2** | | | |
| 0 | 459 (85.6%) | 344 (85.6%) | 115 (85.8%) |
| 1 | 77 (14.4%) | 58 (14.4%) | 19 (14.2%) |

| | | | |
|---|---|---|---|
| N/A | - | - | - |
| **PR Nodal 2** | | | |
| 0 | 279 (52.1%) | 211 (52.5%) | 68 (50.7%) |
| 1 | 257 (47.9%) | 191 (47.5%) | 66 (49.3%) |
| N/A | - | - | - |
| **SD Primary 2** | | | |
| 0 | 534 (99.6%) | 400 (99.5%) | 134 (100.0%) |
| 1 | 2 (0.4%) | 2 (0.5%) | - |
| N/A | - | - | - |
| **SD Nodal 2** | | | |
| 0 | 526 (98.1%) | 396 (98.5%) | 130 (97.0%) |
| 1 | 10 (1.9%) | 6 (1.5%) | 4 (3.0%) |
| N/A | - | - | - |
| **DLT - Dermatological 2** | | | |
| No | 522 (97.4%) | 392 (97,5%) | 130 (97.0%) |
| Yes | 14 (2.6%) | 10 (2.5%) | 4 (3.0%) |
| **DLT - Neurological 2** | | | |
| No | 516 (96.3%) | 386 (96.0%) | 130 (97.0%) |
| Yes | 20 (3.7%) | 16 (4.0%) | 4 (3.0%) |
| **DLT - Gastrointestinal 2** | | | |
| No | 508 (94.8%) | 380 (94.5%) | 128 (95.5%) |
| Yes | 28 (5.2%) | 22 (5.5%) | 6 (4.5%) |
| **DLT - Hematological 2** | | | |
| No | 512 (95.5%) | 382 (95.0%) | 130 (97.0%) |
| Yes | 24 (4.5%) | 20 (5.0%) | 4 (3.0%) |

| | | | |
|---|---|---|---|
| **DLT - Nephrological 2** | | | |
| No | 529 (98.7%) | 397 (98.8%) | 132 (98.5%) |
| Yes | 7 (1.3%) | 5 (1.2%) | 2 (1.5%) |
| **DLT - Vascular 2** | | | |
| No | 535 (99.8%) | 401 (99.8%) | 134 (100.0%) |
| Yes | 1 (0.2%) | 1 (0.2%) | - |
| **DLT - Other 2** | | | |
| No | 522 (97.4%) | 390 (97.0%) | 132 (98.5%) |
| Yes | 14 (2.6%) | 12 (3.0%) | 2 (1.5%) |
| **Decisions** | | | |
| **Decision 1 (Induction Chemo) Y/N** | | | |
| No | 342 (63.8%) | 250 (62.2%) | 92 (68.7%) |
| Yes | 194 (36.2%) | 152 (37.8%) | 42 (31.3%) |
| **Decision 2 (CC / RT alone)** | | | |
| CC | 410 (76.5%) | 313 (77.9%) | 97 (72.4%) |
| RT alone | 126 (23.5%) | 89 (22.1%) | 37 (27.6%) |
| **Decision 3 Neck Dissection (Y/N)** | | | |
| No | 425 (79.3%) | 318 (79.1%) | 107 (79.9%) |
| Yes | 111 (20.7%) | 84 (20.9%) | 27 (20.1%) |
| **Final Outcomes (after D3)** | | | |
| **Overall Survival (4 Years)** | | | |
| Alive | 457 (85.3%) | 344 (85.6%) | 113 (84.3%) |
| Dead | 79 (14.7%) | 58 (14.4%) | 21 (15.7%) |
| N/A | - | - | - |
| **Feeding tube 6 months** | | | |

| | | | |
|---|---|---|---|
| No | 438 (81.7%) | 325 (80.8%) | 113 (84.3%) |
| Yes | 98 (18.3%) | 77 (19.2%) | 21 (15.7%) |
| **Aspiration rate Post-therapy** | | | |
| No | 438 (81.7%) | 323 (80.3%) | 115 (85.8%) |
| Yes | 98 (18.3%) | 79 (19.7%) | 19 (14.2%) |
| **Dysphagia** | | | |
| No | 382 (71.3%) | 280 (69.7%) | 102 (76.1%) |
| Yes | 154 (28.7%) | 122 (30.3%) | 32 (23.9%) |

**eTable 2: Absolute improvement over physicians' results with treatment simulation, per model, without and with radiomics, and for different model outcomes (OS vs. OS+DP).**

In brackets are the 95% confidence intervals. Best results are shown in bold.

| *Model Outcome* | *Radiomics* | *Hidden Layers* | *Training* | | *Testing* | |
|---|---|---|---|---|---|---|
| | | | *Overall Survival (4 Years)* | *Dysphagia* | *Overall Survival (4 Years)* | *Dysphagia* |
| Overall Survival | No | 0 | -0.25% (-14.43%, +6.97%) | | +0.75% (-12.69%, +8.96%) | |
| | | 1 | +3.73% (-6.23%, +8.21%) | | +4.48% (-5.22%, +10.45%) | |
| | | 2 | +2.24% (-9.45%, +7.96%) | | +2.99% (-9.7%, +9.7%) | |
| | | 3 | +3.73% (-2.99%, +8.71%) | | +4.48% (-2.26%, +9.7%) | |
| | | 4 | +1.49% (-2.24%, +8.96%) | | +2.24% (-1.49%, +10.47%) | |
| | | 5 | +1.74% (-4.23%, **+9.2%**) | | +2.99% (-3.73%, +10.45%) | |
| | | 6 | +1.49% (-7.71%, +8.46%) | | +2.99% (-6.72%, **+11.19%**) | |
| | | 7 | +1.49% (-12.44%, +8.46%) | | +2.99% (-9.72%, +10.45%) | |

| | | | | | | |
|---|---|---|---|---|---|---|
| | | 8 | +1.0% (-17.67%, +8.46%) | | +2.24% (-14.93%, +10.45%) | |
| | Yes | 0 | -0.5% (-12.46%, +6.22%) | | -0.75% (-13.43%, +8.23%) | |
| | | 1 | +2.74% (-4.98%, +7.46%) | | +3.73% (-5.97%, +9.7%) | |
| | | 2 | +2.99% (-2.25%, +7.46%) | | +3.73% (-2.99%, +8.96%) | |
| | | 3 | +1.74% (-1.99%, +6.97%) | | +3.73% (-1.49%, +9.7%) | |
| | | 4 | +1.49% (-2.49%, +7.71%) | | +3.73% (-1.49%, +10.45%) | |
| | | 5 | +1.49% (-3.73%, +8.46%) | | +3.73% (-4.48%, **+11.19%**) | |
| | | 6 | +1.74% (-6.22%, +7.96%) | | +2.99% (-7.48%, **+11.19%**) | |
| | | 7 | +1.49% (-10.71%, +7.71%) | | +2.99% (-12.69%, +10.45%) | |
| | | 8 | +0.75% (-12.94%, +7.46%) | | +1.49% (-13.43%, +10.45%) | |
| | | 9 | +0.5% (-15.93%, +7.21%) | | +1.49% (-16.42%, +9.7%) | |
| Overall Survival and Dysphagia | No | 0 | +3.73% (-5.47%, +8.46%) | **+5.97%** (-4.73%, +12.44%) | +4.48% (-5.22%, +10.45%) | +0.75% (-13.43%, **+8.97%**) |
| | | 1 | +3.73% (-1.24%, +7.96%) | +4.98% (-1.75%, +11.19%) | +4.48% (-1.49%, +9.7%) | **+1.49%** (-7.46%, +8.23%) |

| | | | | | | |
|---|---|---|---|---|---|---|
| | | 2 | +2.74% (**-0.25%**, +6.47%) | +3.98% (**-1.24%**, +9.2%) | +3.73% (**-0.75%**, +8.96%) | +0.75% (**-4.48%**, +6.72%) |
| | | 3 | +1.74% (-1.24%, +6.47%) | +1.99% (-2.74%, +8.96%) | +2.99% (**-0.75%**, +8.21%) | -0.75% (**-4.48%**, +5.97%) |
| | | 4 | +1.0% (-1.74%, +8.21%) | +1.24% (-2.99%, +11.95%) | +2.24% (-1.49%, +9.7%) | -1.49% (**-4.48%**, +7.46%) |
| | | 5 | +1.49% (-1.99%, +8.96%) | +1.74% (-3.98%, +12.94%) | +2.99% (-1.49%, **+11.19%**) | -0.75% (-7.46%, +8.21%) |
| | | 6 | +2.24% (-2.99%, +8.96%) | +1.99% (-7.71%, +12.94%) | +3.73% (-2.99%, **+11.19%**) | -0.75% (-15.67%, +8.21%) |
| | | 7 | +2.24% (-7.72%, +8.96%) | +2.24% (-11.69%, **+13.43%**) | +3.73% (-5.99%, +10.47%) | -1.49% (-20.15%, **+8.97%**) |
| | | 8 | +1.74% (-14.43%, +8.46%) | +1.24% (-12.2%, +12.69%) | +2.99% (-12.69%, +10.45%) | -2.99% (-22.39%, +8.96%) |
| | Yes | 0 | -0.5% (-12.2%, +5.72%) | -6.97% (-17.91%, +1.99%) | 0.0% (-13.43%, +8.21%) | -13.43% (-27.63%, -2.24%) |
| | | 1 | +2.74% (-5.47%, +7.21%) | -2.74% (-11.95%, +5.22%) | +3.73% (-5.22%, +9.7%) | -6.72% (-19.4%, +2.24%) |
| | | 2 | **+4.23%** (-2.0%, +7.96%) | 0.0% (-8.46%, +6.22%) | **+5.22%** (-2.26%, +10.45%) | -3.73% (-14.18%, +2.99%) |
| | | 3 | +3.73% (-1.24%, +7.47%) | -0.25% (-7.96%, +6.47%) | **+5.22%** (**-0.75%**, +9.7%) | -2.99% (-12 .71%, +3.73%) |
| | | 4 | +1.0% (-2.24%, +8.21%) | -4.23% (-11.19%, +4.73%) | +2.99% (**-0.75%**, **+11.19%**) | -4.48% (-15.67%, +1.49%) |
| | | 5 | +1.74% (-3.73%, +8.21%) | -4.48% (-20.15%, +5.73%) | +3.73% (-4.5%, +10.45%) | -5.22% (-26.88%, +1.49%) |

| | | | | | | |
|---|---|---|---|---|---|---|
| | | 6 | +1.74% (-6.98%, +7.96%) | -4.98% (-22.39%, +6.72%) | +3.73% (-8.96%, **+11.19%**) | -7.46% (-32.84%, +2.99%) |
| | | 7 | +1.24% (-11.2%, +7.46%) | -6.09% (-24.38%, +6.47%) | +2.24% (-11.96%, +10.45%) | -10.45% (-35.82%, +2.24%) |
| | | 8 | +0.75% (-14.43%, +7.46%) | -5.97% (-24.14%, +6.22%) | +1.49% (-15.67%, +10.45%) | -11.19% (-35.07%, +2.24%) |
| | | 9 | 0.0% (-17.16%, +7.21%) | -7.21% (-25.63%, +6.72%) | +0.75% (-16.44%, +9.7%) | -13.43% (-37.31%, +2.99%) |

**eTable 3: Model similarity to physicians' decisions on training and testing data, without radiomics and with radiomics, and for different model outcomes (OS and OS+DP).**

In brackets are the 95% confidence intervals. Best results are shown in bold.

| *Mode Outcome* | *Radiomics* | *Hidden Layers* | *Training* | | | | *Testing* | | | |
|---|---|---|---|---|---|---|---|---|---|---|
| | | | *Decision 1 (Induction Chemo) Y/N* | *Decision 2 (CC / RT alone)* | *Decision 3 Neck Dissection (Y/N)* | *<u>Overall</u>* | *Decision 1 (Induction Chemo) Y/N* | *Decision 2 (CC / RT alone)* | *Decision 3 Neck Dissection (Y/N)* | *<u>Overall</u>* |
| Overall Survival | No | 0 | 53.23% (39.79%, 63.68%) | 51.99% (34.58%, 67.41%) | 53.98% (38.81%, 67.91%) | 52.65% (43.78%, 60.62%) | 53.73% (37.29%, 68.66%) | 52.24% (35.07%, 69.4%) | 53.73% (34.33%, 70.15%) | 52.99% (42.52%, 61.95%) |
| | | 1 | 52.74% (39.79%, 63.68%) | 52.24% (35.81%, 66.67%) | 71.64% (55.47%, 78.11%) | 58.37% (50.58%, 65.34%) | 53.73% (38.06%, 67.18%) | 51.49% (35.8%, 67.16%) | 71.64% (52.99%, 79.1%) | 58.46% (49.0%, 66.42%) |
| | | 2 | 55.47% (42.53%, 64.93%) | 56.72% (37.81%, 70.9%) | 67.16% (47.01%, 77.61%) | 59.37% (49.59%, 66.92%) | 56.72% (39.55%, 68.66%) | 55.97% (38.04%, 70.9%) | 66.42% (44.76%, 77.63%) | 58.96% (48.5%, 67.66%) |
| | | 3 | 56.72% (43.28%, 65.17%) | 59.95% (41.29%, 73.88%) | 77.61% (61.44%, 79.35%) | 64.1% (55.31%, 70.07%) | 57.46% (40.3%, 68.68%) | 58.96% (40.3%, 71.64%) | 77.61% (61.19%, **80.6%**) | 63.68% (54.73%, 70.9%) |

| | | | | | | | | | | |
|---|---|---|---|---|---|---|---|---|---|---|
| | | 4 | 58.96% (39.3%, 65.17%) | 74.88% (46.27%, **78.11%**) | 78.61% (63.18%, **79.6%**) | 69.49% (59.12%, 73.47%) | 61.19% (36.57%, 69.4%) | 70.9% (47.76%, 73.88%) | 79.1% (62.67%, 79.85%) | 68.91% (57.71%, 73.38%) |
| | | 5 | 56.72% (38.56%, 64.68%) | 74.38% (39.28%, **78.11%**) | 77.11% (47.75%, **79.6%**) | 67.33% (53.56%, 72.97%) | 58.21% (35.82%, 69.4%) | 70.15% (41.79%, 73.88%) | 77.61% (47.0%, 79.85%) | 66.42% (52.72%, 73.13%) |
| | | 6 | 55.47% (37.56%, 64.43%) | 70.15% (30.09%, 77.86%) | 72.39% (34.33%, 79.1%) | 63.27% (46.01%, 71.64%) | 55.22% (32.84%, 68.68%) | 67.16% (32.84%, 73.88%) | 72.39% (33.58%, 79.85%) | 62.19% (45.77%, 71.89%) |
| | | 7 | 52.99% (37.31%, 63.68%) | 65.05% (26.87%, 77.36%) | 67.16% (28.6%, 78.86%) | 59.08% (42.04%, 70.15%) | 52.99% (33.56%, 68.66%) | 62.69% (29.85%, 73.13%) | 67.16% (28.36%, 79.85%) | 58.46% (41.78%, 70.15%) |
| | | 8 | 51.74% (37.31%, 63.43%) | 59.58% (25.12%, 77.11%) | 61.94% (25.86%, 78.61%) | 55.64% (38.56%, 68.58%) | 51.49% (32.82%, 68.66%) | 58.21% (28.36%, 73.13%) | 61.19% (25.37%, 79.85%) | 55.47% (38.31%, 68.66%) |
| | Yes | 0 | 52.74% (40.3%, 63.68%) | 51.24% (34.58%, 66.67%) | 53.86% (38.56%, 68.16%) | 52.32% (43.94%, 61.12%) | 52.99% (35.82%, 67.91%) | 50.75% (33.56%, 67.91%) | 53.73% (35.07%, 69.4%) | 52.49% (42.29%, 62.69%) |
| | | 1 | 53.98% (41.54%, 63.93%) | 54.98% (37.81%, 69.65%) | 71.89% (55.22%, 78.36%) | 59.78% (51.82%, 66.67%) | 54.48% (38.06%, 67.18%) | 54.48% (37.31%, 68.66%) | 71.64% (52.22%, 79.1%) | 59.7% (50.5%, 67.41%) |

| | | | | | | | | | |
|---|---|---|---|---|---|---|---|---|---|---|
| | | 2 | 57.46% (44.53%, 65.42%) | 62.19% (44.03%, 73.39%) | 76.62% (65.92%, 79.35%) | 64.68% (57.63%, 70.15%) | 58.96% (41.79%, 69.4%) | 60.45% (41.79%, 72.39%) | 76.12% (64.18%, **80.6%**) | 64.43% (56.22%, 70.65%) |
| | | 3 | 58.71% (43.77%, 65.67%) | 71.39% (53.23%, 77.62%) | 78.36% (69.65%, **79.6%**) | 68.82% (61.44%, 72.8%) | 60.45% (41.79%, **70.15%**) | 67.91% (51.49%, **74.63%**) | 78.36% (68.66%, 79.85%) | 68.16% (59.7%, 73.13%) |
| | | 4 | 58.58% (40.05%, 65.17%) | 74.38% (50.49%, 77.86%) | 78.86% (63.17%, **79.6%**) | 69.15% (59.7%, 73.22%) | 60.45% (35.82%, 69.4%) | 70.15% (50.75%, **74.63%**) | 79.1% (64.14%, **80.6%**) | 68.91% (59.45%, 73.39%) |
| | | 5 | 57.21% (38.31%, 64.43%) | 74.38% (37.55%, 77.86%) | 77.36% (50.24%, 79.35%) | 67.21% (54.3%, 72.72%) | 58.96% (34.31%, 69.4%) | 70.15% (41.03%, 73.88%) | 77.61% (48.51%, 79.85%) | 66.67% (52.98%, 72.89%) |
| | | 6 | 54.23% (38.06%, 64.43%) | 70.15% (29.1%, 77.86%) | 73.38% (33.83%, 79.1%) | 63.43% (47.68%, 71.73%) | 55.97% (34.31%, 68.66%) | 66.42% (31.34%, 73.88%) | 73.88% (33.58%, 79.85%) | 62.44% (46.51%, 71.64%) |
| | | 7 | 52.61% (37.31%, 63.43%) | 63.93% (25.62%, 77.36%) | 68.91% (28.6%, 79.1%) | 59.25% (42.54%, 69.82%) | 52.61% (32.82%, 67.91%) | 61.94% (29.83%, 73.13%) | 68.66% (29.09%, 79.85%) | 58.46% (41.79%, 70.15%) |
| | | 8 | 51.99% (37.56%, 63.68%) | 59.33% (24.62%, 77.11%) | 61.19% (23.38%, 78.61%) | 55.64% (37.89%, 68.99%) | 51.49% (32.84%, 68.66%) | 57.46% (28.36%, 73.13%) | 61.94% (23.13%, 79.12%) | 55.22% (38.3%, 69.4%) |

| | | | | | | | | | | |
|---|---|---|---|---|---|---|---|---|---|---|
| | | 9 | 51.99% (37.56%, 63.18%) | 54.98% (24.63%, 77.11%) | 59.2% (24.63%, 78.36%) | 53.98% (36.48%, 68.33%) | 52.24% (32.84%, 68.66%) | 53.73% (28.36%, 73.13%) | 59.7% (24.63%, 79.1%) | 53.98% (36.82%, 68.91%) |
| Overall Survival and Dysphagia | No | 0 | 53.98% (39.55%, 63.93%) | 51.0% (34.58%, 66.92%) | 71.39% (53.98%, 77.86%) | 58.46% (49.59%, 66.17%) | 55.22% (37.31%, 68.66%) | 51.49% (32.84%, 68.66%) | 71.64% (51.47%, 79.1%) | 58.46% (49.0%, 67.41%) |
| | | 1 | 55.47% (42.79%, 65.42%) | 66.17% (48.25%, 75.62%) | 77.86% (67.91%, 79.35%) | 65.88% (58.87%, 70.98%) | 56.72% (38.79%, 69.4%) | 63.43% (45.52%, **74.63%**) | 77.61% (67.16%, **80.6%**) | 65.17% (57.46%, 71.65%) |
| | | 2 | 57.71% (45.27%, **66.42%**) | 75.37% (64.92%, **78.11%**) | **79.1%** (76.11%, 79.35%) | 70.4% (65.34%, 73.63%) | 58.96% (42.52%, 68.66%) | 70.9% (60.45%, **74.63%**) | **79.85%** (75.37%, 79.85%) | 69.65% (63.43%, 73.38%) |
| | | 3 | 59.2% (**47.26%**, 66.17%) | **77.11%** (**68.89%**, **78.11%**) | **79.1%** (**77.36%**, 79.35%) | **71.52%** (**66.5%**, **74.13%**) | 61.19% (**45.5%**, **70.15%**) | **72.39%** (**64.93%**, 73.88%) | **79.85%** (**77.61%**, 79.85%) | **70.65%** (**64.68%**, **73.88%**) |
| | | 4 | **59.95%** (41.29%, 65.67%) | 75.87% (58.7%, **78.11%**) | **79.1%** (**77.36%**, 79.35%) | 71.06% (63.68%, 73.63%) | **61.94%** (37.31%, **70.15%**) | 71.64% (57.44%, **74.63%**) | **79.85%** (76.87%, 79.85%) | 70.4% (62.19%, **73.88%**) |
| | | 5 | 57.71% (39.3%, 65.17%) | 76.12% (50.23%, **78.11%**) | 78.86% (70.88%, 79.35%) | 69.73% (59.62%, 73.55%) | 59.7% (34.33%, 69.4%) | 71.64% (50.73%, 73.88%) | **79.85%** (67.16%, 79.85%) | 69.15% (59.2%, 73.63%) |

| | | | | | | | | | | |
|---|---|---|---|---|---|---|---|---|---|---|
| | | 6 | 56.22% (38.8%, 64.18%) | 72.89% (34.08%, 77.86%) | 77.36% (48.01%, 79.35%) | 66.5% (51.82%, 72.64%) | 58.21% (34.33%, 69.4%) | 68.66% (35.07%, **74.63%**) | 77.61% (47.76%, 79.87%) | 65.67% (50.75%, 72.89%) |
| | | 7 | 53.73% (37.56%, 63.93%) | 67.41% (29.35%, 77.86%) | 73.63% (34.81%, 79.1%) | 62.56% (46.43%, 71.23%) | 54.48% (33.58%, 68.66%) | 64.18% (31.32%, 73.13%) | 73.88% (34.31%, 79.85%) | 61.69% (45.52%, 71.39%) |
| | | 8 | 52.24% (37.56%, 63.69%) | 61.69% (26.12%, 77.11%) | 66.42% (28.6%, 78.86%) | 57.63% (41.29%, 69.49%) | 52.99% (32.84%, 67.91%) | 60.45% (29.1%, 73.13%) | 65.67% (26.87%, 79.85%) | 57.21% (41.04%, 69.65%) |
| | Yes | 0 | 52.99% (40.05%, 64.18%) | 51.49% (35.32%, 66.92%) | 53.73% (38.05%, 67.16%) | 52.57% (44.28%, 60.7%) | 53.73% (35.82%, 67.91%) | 51.49% (34.33%, 67.91%) | 53.73% (35.82%, 68.66%) | 52.24% (42.54%, 61.7%) |
| | | 1 | 53.98% (42.03%, 63.94%) | 55.72% (38.31%, 69.65%) | 71.89% (56.72%, 78.36%) | 59.87% (52.16%, 66.67%) | 54.48% (39.53%, 67.16%) | 54.48% (37.31%, 69.4%) | 71.64% (54.48%, 79.1%) | 59.7% (51.24%, 67.41%) |
| | | 2 | 54.23% (42.54%, 64.43%) | 53.36% (36.56%, 69.65%) | 76.37% (64.66%, 79.35%) | 60.95% (53.65%, 68.0%) | 54.48% (39.55%, 67.91%) | 52.99% (35.07%, 68.66%) | 76.12% (62.69%, **80.6%**) | 60.7% (52.24%, 68.41%) |
| | | 3 | 56.22% (43.53%, 65.17%) | 62.44% (43.77%, 74.88%) | 78.86% (70.89%, 79.35%) | 65.34% (58.37%, 70.48%) | 56.72% (40.3%, 68.66%) | 59.7% (41.79%, 72.39%) | **79.85%** (70.88%, **80.6%**) | 64.93% (57.46%, 71.14%) |

| | | | | | | | | | | |
|---|---|---|---|---|---|---|---|---|---|---|
| | | 4 | 59.2% (40.29%, 65.17%) | 75.62% (55.21%, **78.11%**) | 78.86% (62.44%, 79.36%) | 70.07% (59.95%, 73.38%) | **61.94%** (36.57%, 69.42%) | 70.9% (52.97%, 73.88%) | 79.1% (61.18%, 79.85%) | 69.4% (58.71%, 73.64%) |
| | | 5 | 57.21% (38.06%, 64.43%) | 74.38% (36.07%, **78.11%**) | 77.61% (45.01%, 79.35%) | 67.0% (52.9%, 72.89%) | 59.7% (34.33%, 69.42%) | 70.15% (38.06%, 73.88%) | 77.61% (44.03%, 79.85%) | 66.17% (52.73%, 73.13%) |
| | | 6 | 54.48% (38.06%, 64.18%) | 69.65% (28.59%, 77.86%) | 73.13% (33.33%, 79.1%) | 63.39% (46.68%, 71.89%) | 55.97% (33.58%, 68.66%) | 66.42% (32.09%, 73.88%) | 73.13% (32.09%, 79.85%) | 62.69% (46.27%, 71.89%) |
| | | 7 | 52.49% (37.31%, 63.18%) | 63.93% (26.11%, 77.12%) | 67.04% (30.09%, 78.86%) | 58.62% (40.54%, 69.74%) | 52.99% (32.09%, 68.66%) | 61.94% (29.85%, 73.13%) | 67.16% (26.87%, 79.85%) | 58.21% (40.3%, 69.66%) |
| | | 8 | 51.62% (37.81%, 63.43%) | 58.08% (25.61%, 77.11%) | 61.19% (24.87%, 78.61%) | 55.56% (38.56%, 68.41%) | 52.24% (32.84%, 68.66%) | 56.72% (28.36%, 73.13%) | 61.19% (24.63%, 79.1%) | 54.98% (37.81%, 68.41%) |
| | | 9 | 51.0% (37.56%, 63.43%) | 56.22% (24.87%, 77.11%) | 57.09% (24.87%, 78.36%) | 53.32% (37.98%, 67.74%) | 51.49% (32.09%, 68.66%) | 55.22% (28.36%, 72.39%) | 56.72% (23.88%, 79.1%) | 53.23% (37.56%, 67.66%) |

**eMethodology**

One of the state-of-the-art machine learning methods applicable to the optimal therapy process problem is Reinforcement Learning (RL), and in particular Q-Learning. Q-Learning aims to solve problems in which a model has to choose among a series of options to maximize a certain goal in the given situation: the model observes a set of actions and the outcome these actions have, thus learning which choices are optimal and which are not. Q-Learning is thus a type of machine learning, an application of artificial intelligence (AI) that provides systems the ability to automatically learn and improve from experience without being explicitly programmed. Q-Learning has been shown to lead to valid results in a variety of medical problems, be it for the definition of a sequential multiple-assignment randomized trial [1, 2], the optimal treatment of depression [1] and ADHD [2], or to find the breastfeeding habits that maximize child vocabulary development [3].

We use Q-Learning to find a treatment policy that maximizes positive patient outcomes, defined as a linear combination of multiple outcomes, e.g., toxicological and survival outcomes, with different physician specified weights. Specifically, we apply Deep Q-Learning (DQL) to determine the optimal treatment sequence for a curated dataset of 536 HNC patients. The simplified, curated dataset includes three decision points for each patient, related to (1) induction chemotherapy, (2) concurrent chemotherapy or radiotherapy alone, and (3) surgical, i.e., neck dissection, treatment. The resulting fitted models (which vary in depth from linear to 9-layer neural networks) are evaluated on a separate test set of 134 patients, by assessing the similarity to real-life doctors' decisions, as well as by predicting the outcome of the dynamic treatment choices through a separate model built to simulate the effect of treatment decisions based on the patient's history.

**Patient Dataset**

A curated HNC dataset of oropharyngeal squamous cell carcinoma (OPC) patients treated at MD Anderson Cancer Center between 2005 and 2013 was analyzed in this project. All methods for this study were performed in accordance with the University of Texas MD Anderson Cancer Center IRB guidelines and regulations. Being a Health Insurance Portability and Accountability Act (HIPAA)-compliant retrospective study, the prerequisite for informed consent was waived. Clinical features recorded at diagnosis including age at diagnosis, sex, ethnicity, HPV status, smoking status and frequency, subsite of the primary tumor within the oropharynx, T category, N category, therapeutic combination, AJCC stage (8th edition), as well as intermediate results of the treatment decisions, such as Dose-Limiting-Toxicity (DLT) and tumor response, measured between treatment decisions were extracted from electronic medical records. Table 1 shows the demographics of patients for the main clinical features and outcomes considered. Attributes with missing data are also identified in the table. The complete table of clinical demographics is shown in eTable 1.

Tumor response is included as a categorical variable (complete/partial/stable disease for primary tumor and nodal involvement). Furthermore, we also include radiomic features extracted from contrast-enhanced computed tomography (CECT) at initial diagnosis, prior to any active local or systemic treatment. The 3D volumes of interest (VOIs), including the gross primary tumor volumes (GTVp), were manually segmented by a radiation oncologist, and then inspected by a second radiation oncologist within the commercially available contouring software (Velocity AI v3.0.1). The generated VOIs and CT images were exported in the format of DICOM and DICOM-RTSTRUCT to be used for radiomics features extraction. The primary tumor volumes (GTVp) were contoured based on the ICRU 62/83 definition[4]. Radiomics analysis was performed using the freely available open source software "Imaging Biomarker Explorer" (IBEX), which was developed by the University of Texas M.D. Anderson Cancer Center and utilizes the Matlab platform (Mathworks Inc, Natick, VA). The CT images in the format of DICOM and the GTVp contours in the format DICOMRTSTRUCT were imported into IBEX. Extracted features represent the intensity, shape, and texture of the primary tumor.

All the DQL models have been trained with and without the inclusion of the *radiomic* features, to explore if and how their consideration would affect the optimal treatment decisions made by the algorithm. These features are collected at the diagnostic stage.

For each patient, post-treatment outcomes include survival and overall toxicity and toxicity outcomes such as feeding tube and aspiration rate. As a survival outcome we consider Overall Survival at 4 years. No blind assessment of any of the considered outcomes was performed, therefore only patients with enough follow-up time (at least 4 years) or who died before 4 years were considered for the analysis. Among the different outcome measures available, we focus on the following 2 scenarios: a) Considering *Overall Survival (4 Years)* (OS) as a single outcome measure, which refers to whether the patient survived for at least 4 years after the treatment ended; and b) Considering the combination of *Overall Survival (4 years)* and *Dysphagia (DP)* as a multi-outcome measure. In this case, *Dysphagia* is represented by the presence of either *Feeding Tube (FT)* or *Aspiration Rate (AR)* 6 months after the end of the treatment sequence. The combined outcome measure is computed as *OS - (FT + AR PostTherapy - AR PreTherapy).*

For the purpose of this project, we furthermore consider three treatment decision points for each patient as part of the treatment policy: Decision 1 (D1)*: Induction Chemotherapy (IC) or Not*; Decision 2 (D2)*: Concurrent Chemotherapy (CC) or Radiotherapy alone (RT)*; and Decision 3 (D3)*: Neck Dissection (ND).*

**Preprocessing**

The 536 samples of the dataset were split into two distinct sets for training and testing using a 75-25% random split. No blind assessment of the decisions or outcomes was made. Unknown HPV status was handled using a distinguished value (0). Missing values for all other covariates were handled using single imputation: median for numerical variables, mode for categorical ones. The ordinal covariates pathological grade, T-category, N-category, AJCC, prescribed chemo (none/single/doublet/triplet/quadruplet) were coded as numerical features. Given the relatively small sample size, and to reduce dimensionality of the radiomic features (~1000), we applied Principal Component Analysis (PCA) and kept the 6 top components, which explain 90% of the overall variance of the features. After these preprocessing steps, all features were rescaled in the [-1,+1] range, as is standard procedure when training neural networks.

**Q-Learning**

Reinforcement learning (RL) methods, and Q-learning in particular, aim to solve problems in which a model has to choose from a set of possible actions the action that maximizes a certain numerical goal in a given situation. RL models do not learn to replicate the actions performed by other agents (in this case, the physicians' treatment decisions), but rather they observe a sequence of states, the actions that were performed (either by external agents or by the models themselves) in those states, and the outcome of these actions. Based on these observations, RL models learn which actions yield the maximum reward.

The Q-Learning algorithm is a type of RL that learns a Q-function, which, given a state and an action, assigns a value representing the desirability of performing that action in the given state.

Intuitively, a higher Q-function value means that the action is more likely to lead to a positive outcome in the given state. The optimal treatment is then determined as the treatment option with the highest associated Q-value. Since the inputs of Q-functions involve continuously-valued states, conventional Q-learning resorted to approximating the Q-function by a linear function of the state. However, this turns out to be overly simplistic in general, and the recent development of Deep Q-learning (DQL) significantly improved the performance by using neural networks (NN) as the Q-function, turning the task of learning the Q-function into learning the parameters in the network [5].

Traditional Q-learning algorithms require that the models interact with the environment during training, applying a trial-and-error strategy. This is of course not feasible in a dynamic treatment problem, as it would involve the treatment of real people following the directions of a not yet optimal model, which would be problematic both for ethical reasons and time constraints (if the model needs to treat each patient from beginning to post-treatment follow-up it would take decades for the model to be completely trained).

Our models were constructed following the same Q-learning algorithm presented by Moodie et al. [3], in which allows the models to learn from observational data by revisiting the Reinforcement Learning problem as a Supervised Learning problem: for the last treatment decision, the Q-function is the function that maps history and final decision to the final outcome (either Overall Survival or the combined outcome as specified above): the RL problem is thus translated to learning the relationship between patient history, treatment decision. The training is then carried out as in any other regression problem, with a dataset of patients and backpropagation between each epoch, until convergence. There is no need for episodes or rewards, as would be the case with an interactive RL algorithm. For the intermediate decisions, the training is done recursively by using as the variable to predict the Q value computed by the model of the following decision.

For this work, we implemented DQL to model the dynamic decision process at the 3 distinct treatment decision points. eFigure 1a) shows an overview of the training process. A separate neural network model was trained for each of the 3 decision points (D1-D3, respectively), each model being constructed recursively based on the results of the model representing the subsequent decision point (or the outcomes in the case of D3). The features included at each decision point represent the complete history of the patient up to the current treatment decision: D1 only considers variables available at diagnosis, whereas D2 and D3 also include previous treatment decisions and their outcome (i.e., the tumor response and DLT, the Dose-Limiting-Toxicity).

Our learning approach constructs multiple shallow-to-deep neural networks (with a number of layers varying from 0 to 9), as a linear model would not be able to adequately capture the problem complexity and optimally solve it. For each category of models, we constructed a series of increasingly deeper NNs, starting from a 0-layer linear model and progressively adding layers, until the deepest model showed poor performance on the training set (due to overfitting). For the models without radiomics, this process resulted in a model range from 0 to 8 hidden layers, whereas models with radiomics generally took longer to overfit, so the range is 0 to 9 hidden layers. To estimate the statistics of our results, we sampled 1000 separate training sets from the initial training data, and trained a separate model on each of these sets, thus obtaining bootstrapped models. This bootstrapping allows us to give more precise results by providing 95% confidence intervals.

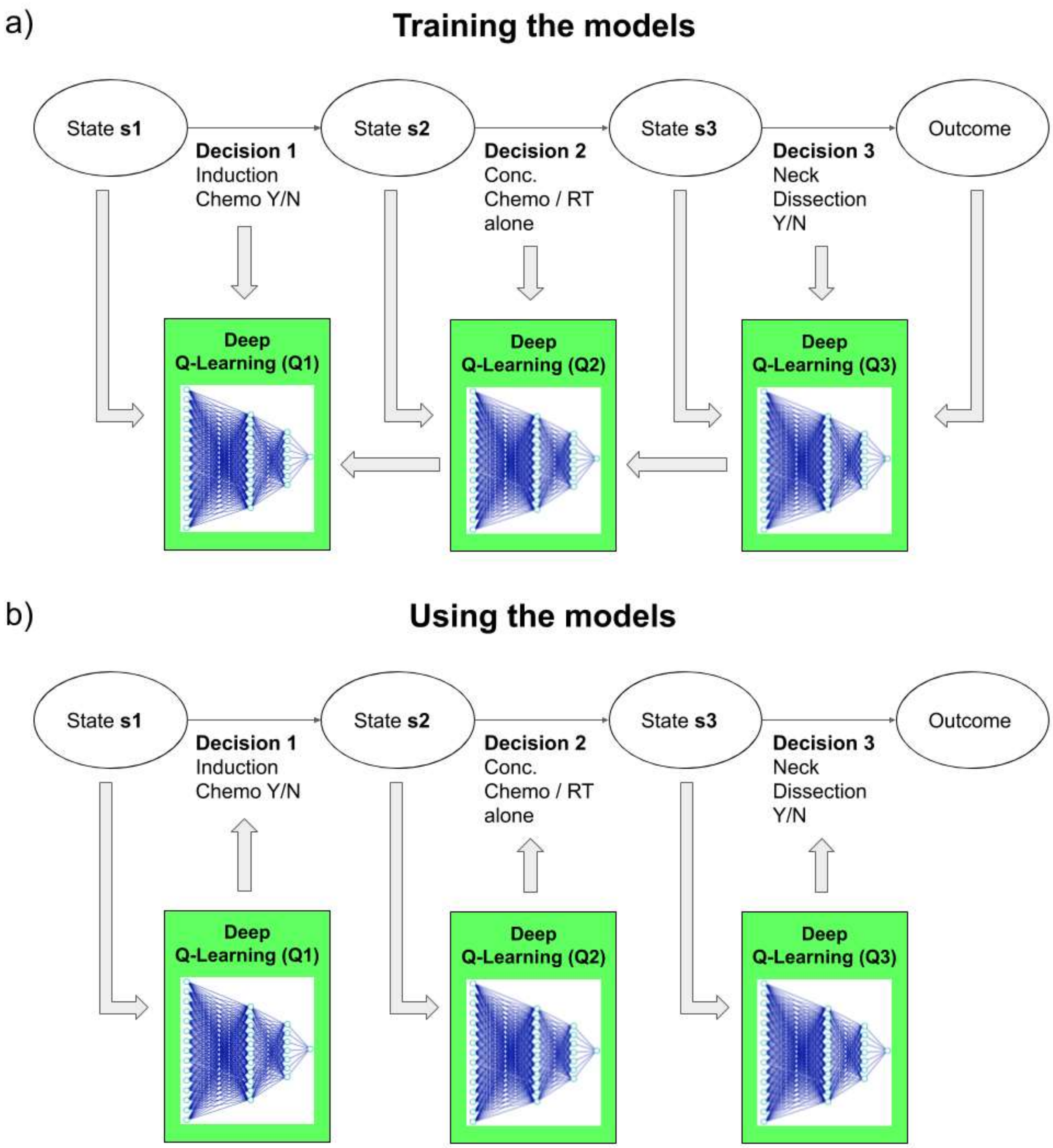


**eFigure 1. Overview of DQL model training and usage.**

a) Overview of DQL model training. The first model to be trained is Q3, which represents *Neck Dissection* (D3), based on the final outcomes, the treatment decisions made in D3, and the patient's history before D3. The model for D2 is then trained based on the result of Q3 instead of the final outcomes, and D1 based on the result of Q2. Once the models are trained, they are used in forward order, opposite to the training order, to prescribe the optimal treatment at each decision step. b) Overview of using the DQL models to prescribe optimal treatment. Once the models are trained, they are used in forward order, opposite to the training order, to prescribe the optimal treatment at each decision step.

eFigure 1b) shows how the trained models are used to prescribe the optimal treatment. By prescribing an optimal treatment at each treatment junction, the DQL models construct a *digital twin* of the decision process, even though their objective is not to replicate the physicians' decisions.

The models have been constructed and trained using the PyTorch framework with GPU acceleration. We report training times in the Results section.

**Treatment Simulator and Evaluation**

Because the goal of DQL is not to replicate doctors' decisions, but to find an optimal treatment, which may differ from what the physician prescribed, our evaluation includes building a separate model which, given a patient's history and the prescribed treatment, predicts the outcome of that treatment. This *patient treatment digital twin* approach enables us to simulate the results of applying the Q-Learning models to patients, and compare the outcomes to the repository recorded outcomes resulting from the clinicians' decisions.

To this end, we built a Treatment Simulator (TS), which, based on the patient's history and treatment decisions, predicts the outcome at the next step. This simulator contains a separate model for each intermediate and final outcome measure, built using a Support Vector Classifier (SVC) and tuned via 5-fold cross-validation over the training data to select the best kernel (between gaussian, sigmoid, or polynomial) and hyperparameters. These models serve as an *in silico digital twin* of the patient treatment, as we can use them to dynamically simulate the patient's *in vivo* course as a function given treatment policy, without physically having to treat the patient.

The prediction accuracy of the TS with 95% confidence intervals was assessed with out of bag evaluation of 1000 models trained on stratified bootstrapped samples. The full details of the SVC are provided in the eTable 4.

We note that although many RL algorithms learn by applying the policy in an environment, we do not use the TS in the training phase, and only limit its use to evaluation. This approach, based on learning from observation data, enables us to avoid the risk of overfitting the DQL models to the simulated environment. This results in a more objective evaluation of the learned DQL models.

The TS performance was also evaluated at the whole policy level by comparing its final outcome prediction against the recorded final outcomes, when the TS followed the same policy as the one prescribed by the physicians, for each patient in the test dataset.

Using the TS, we can then compare the outcomes yielded by the DQL decisions with those yielded by the physicians' prescriptions, thus evaluating the DQL effectiveness. Furthermore, to objectively compare the DQL decisions to the physicians' decisions and facilitate interpretation, we compute the similarity between each DQL model's decisions and the physicians' decisions, considering each decision point independently, and without inclusion of the treatment simulator (TS). To this end, at each decision point, we compute the percentage of patients (train or test) for which the DQL model made the same decision as the physicians at that decision point, regardless of what the previous decision(s) were. An overall similarity score is computed by averaging the similarity for each decision point for each model.

We also evaluated the treatment decisions made by the Q-learning models by examining compliance with the NCCN guidelines of acceptable care [6] which state that eligible advanced stage patients (T3-4 or N1-2-3) must be prescribed chemotherapy, either induction (D1) or concurrent (D2). It is worth noting, that these guidelines/restrictions were not explicitly introduced during model training.

To further support interpretation, the general policy followed by each of the Q-learning models was analyzed by computing the increase (or decrease) in prescription rate for each treatment decision compared to the physicians' *ad-hoc* prescriptions, to express whether each model is more (or less) likely to prescribe a certain treatment when compared to the actual physician.

All evaluations have been performed on a separate test set of 134 patients.

**etable 4: SVC details**

Detailed training parameters of each SVC model, without and with Radiomics: kernel type, C value, degree (only valid for polynomial kernel), gamma value, and class weight.

| *Radiomics* | *Outcome* | *C* | *Kernel* | *Degree* | *Gamma* | *Class Weight* |
|---|---|---|---|---|---|---|
| No | Prescribed Chemo (Single/doublet/triplet/quadruplet/none/NOS) | 3 | polynomial | 5 | automatic | balanced |

| | | | | | | |
|---|---|---|---|---|---|---|
| | Chemo Modification (Y/N) | 1 | gaussian | - | automatic | balanced |
| | Dose modified | 4 | polynomial | 14 | automatic | balanced |
| | Dose delayed | 60 | gaussian | - | automatic | balanced |
| | Dose cancelled | 11 | gaussian | - | automatic | balanced |
| | Regimen modification | 3 | polynomial | 20 | automatic | balanced |
| | DLT (Y/N) | 2 | gaussian | - | automatic | balanced |
| | DLT_Dermatological | 2 | polynomial | 15 | automatic | balanced |
| | DLT_Neurological | 150 | gaussian | - | automatic | balanced |
| | DLT_Gastrointestinal | 400 | polynomial | 3 | automatic | balanced |
| | DLT_Hematological | 300 | gaussian | - | automatic | balanced |
| | DLT_Nephrological | 2 | polynomial | 20 | automatic | balanced |
| | DLT_Vascular | 2 | gaussian | - | automatic | balanced |
| | DLT_Infection (Pneumonia) | 1 | polynomial | 15 | automatic | balanced |
| | DLT_Other | 3 | gaussian | - | automatic | balanced |
| | DLT_Grade | 1 | polynomial | 10 | automatic | balanced |
| | No imaging (0=N, 1=Y) | 1 | gaussian | - | automatic | balanced |
| | CR Primary | 1000 | gaussian | - | automatic | balanced |
| | CR Nodal | 2 | polynomial | 15 | automatic | balanced |
| | PR Primary | 2 | gaussian | - | automatic | balanced |
| | PR Nodal | 1 | gaussian | - | automatic | balanced |
| | SD Primary | 10 | gaussian | - | automatic | balanced |
| | SD Nodal | 3 | polynomial | 15 | automatic | balanced |
| | CC Regimen | 10000 | gaussian | - | automatic | balanced |
| | CC modification (Y/N) | 10000 | gaussian | - | automatic | balanced |
| | CR Primary 2 | 2 | polynomial | 30 | automatic | balanced |
| | CR Nodal 2 | 3 | sigmoid | - | automatic | balanced |
| | PR Primary 2 | 10000 | polynomial | 20 | automatic | balanced |
| | PR Nodal 2 | 10000 | polynomial | 50 | automatic | balanced |
| | SD Primary 2 | 1 | gaussian | - | automatic | balanced |
| | SD Nodal 2 | 10 | gaussian | - | automatic | balanced |

| | | | | | | |
|---|---|---|---|---|---|---|
| | DLT_Dermatological 2 | 100 | gaussian | - | automatic | balanced |
| | DLT_Neurological 2 | 10 | polynomial | 50 | automatic | balanced |
| | DLT_Gastrointestinal 2 | 30 | polynomial | 30 | automatic | balanced |
| | DLT_Hematological 2 | 100 | polynomial | 20 | automatic | balanced |
| | DLT_Nephrological 2 | 3 | polynomial | 3 | automatic | balanced |
| | DLT_Vascular 2 | 1 | gaussian | - | automatic | balanced |
| | DLT_Other 2 | 2 | polynomial | 30 | automatic | balanced |
| | Overall Survival (4 Years) | 100 | gaussian | - | automatic | balanced |
| | Feeding tube 6m | 10 | gaussian | - | automatic | balanced |
| | Aspiration rate Post-therapy | 10 | gaussian | - | automatic | balanced |
| Yes | Prescribed Chemo (Single/doublet/triplet/quadruplet/none/NOS) | 5 | gaussian | - | automatic | balanced |
| | Chemo Modification (Y/N) | 5 | gaussian | - | automatic | balanced |
| | Dose modified | 100 | polynomial | 20 | automatic | balanced |
| | Dose delayed | 100 | polynomial | 10 | automatic | balanced |
| | Dose cancelled | 100 | gaussian | - | automatic | balanced |
| | Regimen modification | 10 | polynomial | 15 | automatic | balanced |
| | DLT (Y/N) | 10 | polynomial | 10 | automatic | balanced |
| | DLT_Dermatological | 500 | gaussian | - | automatic | balanced |
| | DLT_Neurological | 100 | gaussian | - | automatic | balanced |
| | DLT_Gastrointestinal | 200 | polynomial | 10 | automatic | balanced |
| | DLT_Hematological | 10000 | polynomial | 10 | automatic | balanced |
| | DLT_Nephrological | 1000 | polynomial | 10 | automatic | balanced |
| | DLT_Vascular | 4 | gaussian | - | automatic | balanced |
| | DLT_Infection (Pneumonia) | 1000 | gaussian | - | automatic | balanced |
| | DLT_Other | 1 | gaussian | - | automatic | balanced |

| | | | | | | |
|---|---|---|---|---|---|---|
| | DLT_Grade | 1000 | polynomial | 15 | automatic | balanced |
| | No imaging (0=N, 1=Y) | 1 | gaussian | - | automatic | balanced |
| | CR Primary | 100 | gaussian | - | automatic | balanced |
| | CR Nodal | 10 | polynomial | 14 | automatic | balanced |
| | PR Primary | 1 | gaussian | - | automatic | balanced |
| | PR Nodal | 1 | gaussian | - | automatic | balanced |
| | SD Primary | 4 | polynomial | 10 | automatic | balanced |
| | SD Nodal | 1000 | polynomial | 5 | automatic | balanced |
| | CC Regimen | 1000 | polynomial | 20 | automatic | balanced |
| | CC modification (Y/N) | 10000 | gaussian | - | automatic | balanced |
| | CR Primary 2 | 5 | polynomial | 20 | automatic | balanced |
| | CR Nodal 2 | 3 | sigmoid | - | automatic | balanced |
| | PR Primary 2 | 100000 | polynomial | 60 | automatic | balanced |
| | PR Nodal 2 | 10000 | polynomial | 30 | automatic | balanced |
| | SD Primary 2 | 1 | gaussian | - | automatic | balanced |
| | SD Nodal 2 | 100 | gaussian | - | automatic | balanced |
| | DLT_Dermatological 2 | 1000 | polynomial | 30 | automatic | balanced |
| | DLT_Neurological 2 | 1000 | gaussian | - | automatic | balanced |
| | DLT_Gastrointestinal 2 | 10000 | polynomial | 10 | automatic | balanced |
| | DLT_Hematological 2 | 1000 | polynomial | 30 | automatic | balanced |
| | DLT_Nephrological 2 | 10 | gaussian | - | automatic | balanced |
| | DLT_Vascular 2 | 1 | gaussian | - | automatic | balanced |
| | DLT_Other 2 | 100 | gaussian | - | automatic | balanced |
| | Overall Survival (4 Years) | 100 | gaussian | - | automatic | balanced |
| | Feeding tube 6m | 10 | gaussian | - | automatic | balanced |
| | Aspiration rate Post-therapy | 4 | gaussian | - | automatic | balanced |

**eTable 5: TRIPOD checklist**

This checklist is adapted from the Tripod checklist for predictive ML models, taking into account that our model is not a predictive one, but one that seeks to optimize a treatment sequence.

| Section/Topic | Item | Checklist item | Page |
|---|---|---|---|
| **Title and abstract** | | | |
| Title | 1 | Identify the study as developing the model, the target population, and the model purpose | 1 |
| Abstract | 2 | Provide a summary of objectives, study design, setting, participants, sample size, variables, outcome, statistical analysis, results, and conclusions. | 3-4 |
| **Introduction** | | | |
| Background and objectives | 3a | Explain the medical context and rationale for developing or validating the model, including references to existing models. | 5-6 |
| | 3b | Specify the objectives, including whether the study describes the development or | 6 |

| | | | |
|---|---|---|---|
| | | validation of the model or both. | |
| **Methods** | | | |
| Source of data | 4a | Describe the study design or source of data (e.g., randomized trial, cohort, or registry data), separately for the development and validation data sets, if applicable | 7 |
| | 4b | Specify the key study dates, including start of accrual; end of accrual; and, if applicable, end of follow-up. | 7 |
| Participants | 5a | Specify key elements of the study setting (e.g., primary care, secondary care, general population) including number and location of centres. | 7 |
| | 5b | Describe eligibility criteria for participants. | 7 |
| | 5c | Give details of treatments received, if relevant. | 6-7 |
| Outcome | 6a | Clearly define the decisions taken and the outcome maximized by the model, | 6 |

| | | | |
|---|---|---|---|
| | | including how and when assessed. | |
| | 6b | Report any actions to blind assessment of the outcome to be maximized or decisions to be taken. | e16(Supplement) |
| Predictors | 7a | Clearly define all predictors used in developing or validating the multivariable prediction model, including how and when they were measured. | 7, 22-27 (Main manuscript)<br>e2-e8 (Supplement) |
| | 7b | Report any actions to blind assessment of features. | e14 (Supplement) |
| Sample size | 8 | Explain how the study size was arrived at | 7 |
| Missing data | 9 | Describe how missing data were handled (e.g., complete-case analysis, single imputation, multiple imputation) with details of any imputation method. | e16 (Supplement) |
| Statistical analysis method | 10a | Describe how variables were handled in the analyses | 7 (Main manuscript)<br>e16 (Supplement) |
| | 10b | Specify type of model, all model-building procedures | 6-9 (Main manuscript)<br>e14-e18 (Supplement) |

| | | | |
|---|---|---|---|
| | | (including any feature selection), and method for internal validation. | |
| | 10d | Specify all measures used to assess model performance and, if relevant, to compare multiple models. | 8-9 (Main manuscript)<br>e18 (Supplement) |
| Risk groups | 11 | Provide details on how risk groups were created, if done. | Not applicable |
| **Results** | | | |
| Participants | 13a | Describe the flow of participants through the study, including the number of participants with and without the outcome and, if applicable, a summary of the follow-up time. A diagram may be helpful. | 7, 9, 22-27 (Main manuscript)<br>e2-e8, e14-e16 (Supplement) |
| | 13b | Describe the characteristics of the participants (basic demographics, clinical features, available predictors), including the number of participants with missing data for predictors and outcome. | 22-27 (Main manuscript)<br>e2-e8 (Supplement) |
| Model development | 14a | Specify the number of | 22-27 (Main manuscript) |

| | | | |
|---|---|---|---|
| | | participants and outcome events in each analysis. | e2-e8 (Supplement) |
| | 14b | If done, report the unadjusted association between each candidate predictor and outcome. | Not applicable. |
| Model specification | 15a | Present the full model to allow usage for individuals (i.e., all regression coefficients, and model intercept or baseline survival at a given time point) | 8 |
| | 15b | Explain how to use the model | 8 (Main manuscript)<br>e17 (Supplement) |
| Model performance | 16 | Report performance measures (with CIs) for the model. | 9-11, 27-28 (Main manuscript)<br>e9-e14 (Supplement) |
| **Discussion** | | | |
| Limitations | 18 | Discuss any limitations of the study (such as non-representative sample, few events per variable, missing data). | 13-14 |
| Interpretation | 19b | Give an overall interpretation of the results, considering objectives, limitations, and results from similar studies, | 11-13 |

| | | | |
|---|---|---|---|
| | | and other relevant evidence. | |
| Implications | 20 | Discuss the potential clinical use of the model and implications for future research | 14 |
| **Other information** | | | |
| Supplementary information | 21 | Provide information about the availability of supplementary resources, such as study protocol, Web calculator, and data sets. | 8 |
| Funding | 22 | Give the source of funding and the role of the funders for the present study. | 15-16 |
